# Photoreceptor layer thinning is an early biomarker for age-related macular degeneration development: Epidemiological and genetic evidence from UK Biobank optical coherence tomography data

**DOI:** 10.1101/2021.08.18.21262226

**Authors:** Seyedeh Maryam Zekavat, Sayuri Sekimitsu, Yixuan Ye, Vineet Raghu, Hongyu Zhao, Tobias Elze, Ayellet V. Segrè, Janey L. Wiggs, Pradeep Natarajan, Lucian Del Priore, Nazlee Zebardast, Jay C. Wang

## Abstract

**Introduction:** Age-related macular degeneration (AMD) is a blinding condition for which there is currently no early-stage clinical biomarker. AMD is characterized by thinning of the outer retina and drusen formation leading to thickening of the Bruch’s membrane and RPE complex, but the timing between these two events, as well as the role of genetic variants in these processes, are unclear. Here, we jointly analyzed genomic, electronic health record, and optical coherence tomography (OCT) data across 44,823 individuals from the UK Biobank to characterize the epidemiological and genetic associations between retinal layer thicknesses and AMD.

**Methods:** The Topcon Advanced Boundary Segmentation algorithm was used for automated retinal layer segmentation. We associated 9 retinal layer thicknesses with prevalent AMD (present at enrollment) in a logistic regression model, and with incident AMD (diagnosed after enrollment) in a Cox proportional hazards model. Next, we tested the association of AMD-associated genetic alleles, individually and as a polygenic risk score (PRS), with retinal layer thicknesses. All analyses were adjusted for age, age^2^, sex, smoking status, and principal components of ancestry.

**Results:** Photoreceptor segment (PS) thinning was observed throughout the lifespan of individuals analyzed and accelerated at age 45, while retinal pigment epithelium and Bruch’s membrane complex (RPE+BM) thickening started after age 57. Each standard deviation (SD) of PS thinning and RPE+BM thickening were associated with prevalent AMD (PS: OR 1.37, 95% CI 1.25-1.49, P=2.5×10^−12^; RPE+BM: OR=1.34, 95% CI 1.27-1.41, P=8.4×10^−28^) and incident AMD (PS: HR 1.35, 95% CI 1.23-1.47, P=3.7×10^−11^; RPE+BM: HR 1.14, 95% CI 1.06-1.22, P=0.00024). An AMD polygenic risk score (PRS) was associated with PS thinning (Beta -0.21 SD per 2-fold genetically increased risk of AMD, 95% CI -0.23 to -0.19, P=2.8×10^−74^), and its association with RPE+BM was U-shaped (thinning with AMD PRS<92^nd^ percentile and thickening with AMD PRS>92^nd^ percentile suggestive of drusen formation). The loci with strongest support were AMD risk-raising variants *CFH*:rs570618-T, *CFH*:10922109-C, and *ARMS2/HTRA1*:rs3750846-C on PS thinning, and *SYN3/TIMP3*:rs5754227-T on RPE+BM thickening.

**Conclusions:** Epidemiologically, PS thinning precedes RPE+BM thickening by decades, and is the retinal layer most strongly predictive of future AMD risk. Genetically, AMD risk variants are associated with decreased PS thickness. Overall, these findings support PS thinning as an early-stage clinical biomarker for future AMD development.

## Introduction

Age-related macular degeneration (AMD) is a complex genetic disease and the leading cause of vision loss for older adults in developed countries, characterized by drusen deposits in the early dry form and neovascularization in the later wet form. Many contributing risk factors exist for AMD, including age, cigarette smoking, and genetics^1^. Furthermore, AMD has been linked to changes in retinal layer thickness on fundoscopy and optical coherence tomography (OCT) imaging that are now widely used for its diagnosis and monitoring. However, despite widespread clinical use of OCT imaging, a large-scale association of distinct retinal layer thicknesses with future AMD risk has yet to be performed. Additionally, for associating layers, whether changes in retinal layers thicknesses occur early enough during the disease pathogenesis to represent a causal pathway towards AMD is also not well understood.

Clinically, one of the earliest signs of AMD detected in fundus examinations is the presence of drusen deposits, composed of proteins, lipids, mucopolysaccharides, and other extracellular debris which appear in between the retinal pigment epithelium (RPE) and Bruch’s membrane (BM) during adulthood and accumulate over time^2^. Prior studies using histology and electron microscopy have also shown that thickening apical to the BM extracellular matrix, in what is known as basal laminar deposits (BLamD)^3,4^, typically precedes the development of drusen. The progressive accumulation of BLamD has been accompanied by poor visual acuity, RPE degeneration, and pigment changes on fundoscopy^5^. Other changes associated with AMD include photoreceptor loss, which has been observed with aging and in both early and advanced AMD^6,7^. The largest prior study of OCT changes in AMD (N=449) identified significant associations with thickening of the RPE+BM and thinning of the PS^8^. While most studies have highlighted the role of the outer retina in AMD, small studies (N∼100) have also suggested that changes in the inner layers are linked to AMD, including thinning of the GCC^9,10^ and IPL^11^ layers; however, these findings were not replicated in the larger study^8^.

AMD is also a complex genetic disease, with overall heritability of 46% from twin studies, and heritability of 71% for advanced AMD^12^. The first genome-wide screen for AMD across 96 cases and 60 controls identified the complement factor H gene, where individuals homozygous for the risk allele had a 7.4-fold (95% CI 2.9-19) increased risk of AMD^13^. Since then, the most-cited large-scale genome-wide association study (GWAS) of AMD across 16,144 patients and 17,832 controls identified 34 unique loci linked to advanced AMD^14^, and estimated genetic heritability to be 46.7% for advanced AMD, similar to that estimated from twin studies. More recently, variants in *CFH* and *ARMS2* loci were not found to be associated with macular thickness averaged within the outermost circle of the Early Treatment of Diabetic Retinopathy Study (ETDRS) map among UK Biobank participants^15^, but a separate study (N=299) found that these loci are associated with perifoveal changes in macular retinal thickness on OCT^16^, suggesting that AMD loci may be associated with local changes in retinal thickness or changes in specific layers of the retina.

Several questions regarding the link between retinal layer thicknesses and AMD remain. First, the timing of drusen buildup and RPE+BM thickening in relation to photoreceptor degeneration remains unclear. Second, a large-scale assessment of the epidemiological associations between different retinal layer thicknesses with both cross-sectional prevalent AMD and future AMD diagnosis has yet to be performed. Third, an unbiased assessment of the association of known AMD genetic variants with different retinal layer thicknesses has yet to be performed. Here, we jointly analyzed genomic, electronic health record, and retinal OCT data from 44,823 Europeans in the UK Biobank to characterize the epidemiological and genetic association between AMD and retinal layer thicknesses derived from OCT imaging.

## Methods

### UK Biobank cohort, AMD phenotype definition, and sample exclusions

The UK Biobank is a population-based cohort of approximately 500,000 participants recruited from 2006-2010 with existing genomic and longitudinal phenotypic data and median 10-year follow-up^17^. Baseline assessments were conducted at 22 assessment centres across the UK with sample collections including blood-derived DNA. Use of the data was approved by the Massachusetts General Hospital Institutional Review Board (protocol 2021P002040) and facilitated through UK Biobank Applications 7089 and 50211.

The AMD phenotype was defined using a combination of main and secondary ICD-10 (Field IDs 41202, 41204: Code H35.3) and ICD-9 (Field IDs 41203, 41205: Code 3625) diagnoses for macular degeneration, self-reported macular degeneration (Field ID 20002: Code 1528), and macular degeneration from the available general practice data. Prevalent AMD cases were defined as individuals who had AMD first diagnosed at or before enrollment. Incident AMD cases were defined as individuals who had AMD first diagnosed after enrollment.

Of the 67,339 genotyped individuals with retinal imaging data from the TOPCON-3D OCT device available at enrollment, we analyzed images across 44,823 European participants consenting to genetic analyses with genotypic-phenotypic sex concordance, and after excluding one from each pair of 1^st^ or 2^nd^ degree relatives selected randomly. Details on removal of poor-quality images, as well as genotyping quality control filters are provided below.

### Retinal optical coherence tomography (OCT) and retinal layer segmentation

Spectral domain OCT scans of the macula were obtained using Topcon 3D OCT 1000 Mk2 (Topcon, Inc, Japan). Three-dimensional macular volume scans were obtained (512 horizontal A-scans/B-scan; 128 B-scans in a 6×6-mm raster pattern)^18^. All OCT images were stored in .fda image files without prior analysis of macular thickness. We used the Topcon Advanced Boundary Segmentation (TABS) algorithm to automatically segment all scans, which uses dual-scale gradient information to allow for automated segmentation of the inner and outer retinal boundaries and retinal sublayers^19^. The boundaries segmented include the internal limiting membrane (ILM), nerve fiber layer (NFL), ganglion cell layer (GCL), inner plexiform layer (IPL), inner nuclear layer (INL), external limiting membrane (ELM), photoreceptor inner segment/outer segment (IS/OS) junction layer, retinal pigment epithelium (RPE), Bruch’s membrane (BM) and choroid-sclera interface (CSI). The software provides an image quality score and segmentation indicators which was used for quality control. Segmentation indicators included the Inner Limiting Membrane (ILM) Indicator, a measure of the minimum localized edge strength around the ILM boundary across the scan, which can be used to identify blinks, scans that contain regions of signal fading, and errors in segmentation^20^. We excluded all images with image quality less than 40 and images representing the poorest 10% of ILM indicator. We also excluded any image with a layer thickness greater than 2.5 standard deviations away from the mean. The thickness of each retinal sub-layer was determined by calculating the difference between boundaries of interest and averaging this across all scans. For example, RNFL thickness was calculated as the difference between ILM and RNFL boundary lines.

### UK Biobank array genotyping and quality control filters

Genome-wide genotyping of blood-derived DNA was performed by UK Biobank across 488,377 individuals using two genotyping arrays sharing 95% of marker content: Applied Biosystems UK BiLEVE Axiom Array (807,411 markers in 49,950 participants) and Applied Biosystems UK Biobank Axiom Array (825,927 markers in 438,427 participants) both by Affymetrix (Santa Clara, CA)^17^. Variants used in the present analysis include those also imputed using the Haplotype Reference Consortium reference panel of up to 39 million bi-allelic variants and 88 million variants from the UK10K+1000 Genomes reference panels)^17^. Poor quality variants and genotypes were filtered as previously described^17^, with additional filters including high-quality imputed variants (INFO score >0.4), minor allele frequency >0.005, and with Hardy-Weinberg Equilibrium P>1×10^−10^, as previously implemented using Hail-0.2 (https://hail.is/docs/0.2/index.html)^21-23^. Across all genetic analyses, we used data for 462,344 European participants consenting to genetic analyses, with genotypic-phenotypic sex concordance, and after excluding one from each pair of 1^st^ or 2^nd^ degree relatives selected randomly.

### AMD polygenic risk score development

A genome-wide AMD PRS was developed using data across 12,023,831 variants from the prior GWAS of 16,144 Advanced AMD cases and 17,832 controls across 26 studies (93% European, 58% female, and with mean age 73 [SD 10y]) published by Fritsche et al. *Nature Genetics* 2016^14^. The PRS was developed using PRS-CS^24^, a Bayesian method that uses the GWAS summary statistics and linkage disequilibrium patterns from an external reference panel to infer the posterior effect size of each variant using a continuous shrinkage (CS) prior. Using the posterior effect sizes of each variant from PRS-CS, plink-2.0’s –score command was then used using the ‘sum’ modifier to combine the posterior effect sizes into a genome-wide score. This AMD PRS was then normalized to mean 0 and SD 1 and scaled such that 1 unit increase in the PRS reflected a 2-fold increased odds of AMD in the UK Biobank; individuals with AMD PRS >1 are in the top 0.7% of the PRS. The final PRS had mean 0, range -1.6 to 2.09, and SD 0.39.

### Epidemiological analyses between retinal layer thickness and prevalent and incident AMD

Epidemiological analyses were performed between normalized retinal layer thicknesses and prevalent AMD using a logistic regression model, and with incident AMD using a Cox Proportional hazards model, adjusted for age, age^2^ (to adjust for non-linear relationships with age), sex, smoking status, and the first ten principal components of genetic ancestry. Further sensitivity analyses were performed adjusting for glaucoma, myopia, type 2 diabetes, and spherical equivalent. Threshold for significance was defined using a Bonferroni threshold given 8 non-overlapping retinal layers as P<0.05/8=0.00625. Analyses were performed using R-3.5.

### Genetic analyses between AMD and retinal layer thickness

Association of the normalized then scaled AMD PRS with normalized retinal layer thickness was performed using a linear regression model adjusted for age, age^2^, sex, smoking status, and the first ten principal components of genetic ancestry. Further sensitivity analysis was performed removing individuals with incident or prevalent AMD, and adjusting for spherical equivalent. Threshold for significance was defined using a Bonferroni threshold given 8 non-overlapping retinal layers as P<0.05/8=0.00625. Additionally, locus-specific analyses were performed assessing the association of 48 independent variants present in the UK Biobank across 34 AMD loci identified in Fritsche et al. Nature Genetics 2016. Linear regression was performed in a model adjusted for age, age^2^, sex, smoking status, and the first ten principal components of genetic ancestry.

## Results

### Baseline characteristics

Across 44,823 unrelated individuals analyzed, mean (standard deviation [SD]) age was 57 (SD 8) years, 24,058 (54%) were female, and 20,490 (46%) either previously or currently smoked (**Supplementary Table 1**). Mean body mass index was 27 (SD 5) kg/m^2^, 951 (2.1%) had prevalent type 2 diabetes mellitus, 2,292 (5.1%) hypertension, 7,452 (16.6%) hypercholesterolemia, and 639 (1.4%) stroke. Prevalence of clinical ocular conditions was low, with 1585 (3.5%) having self-reported or diagnosed history of cataract, 624 (1.4%) glaucoma, 512 AMD (1.1%), and 108 (0.2%) with diagnosed retinal detachment (**Supplementary Table 1**).

### Retinal layer thicknesses

The thickness of 9 separate retinal layers was measured in micrometers (um), including the retinal nerve fiber layer (RNFL), ganglion cell layer (GCL), and inner plexiform layer (IPL), which together makeup the ganglion cell complex (GCC), as well as the inner plexiform layer (IPL), inner nuclear layer (INL), outer plexiform and outer nuclear layer (OPL+ONL), photoreceptor inner and outer segments (PS), retinal pigment epithelium and Bruch’s membrane complex (RPE+BM), and choroid-sclera interface (CSI) (**Figure 1a**). The mean (SD) thickness in um of the GCC was 102.1 (SD 8.2), RNFL 40 (SD 4.7), GCL 33.2 (SD 2.8), IPL 28.9 (SD 2.6), INL 30.9 (SD 2.0), OPL+ONL 77.4 (SD 5.8), PS 63.2 (SD 2.7), RPE+BM 23.2 (SD 1.8), CSI 200.3 (SD 46.1) (**Supplementary Table 1, Figure 1a**). The retinal layer thicknesses were significantly associated with age and sex (**Supplementary Table 2**). Age-associated retinal thinning occurred for all layers except the OPL+ONL which thickened until age 54 years then thinned, and RPE+BM which increased in thickness after age 57 years (**Figure 1b,c**). Of note, PS thinning occurred throughout the age span of individuals studied (40-70 years), however accelerated PS thinning started at age 45 years, 12 years prior to RPE+BM thickening (**Figure 1b,c, Supplementary Figure 1**).

**Figure 1:**
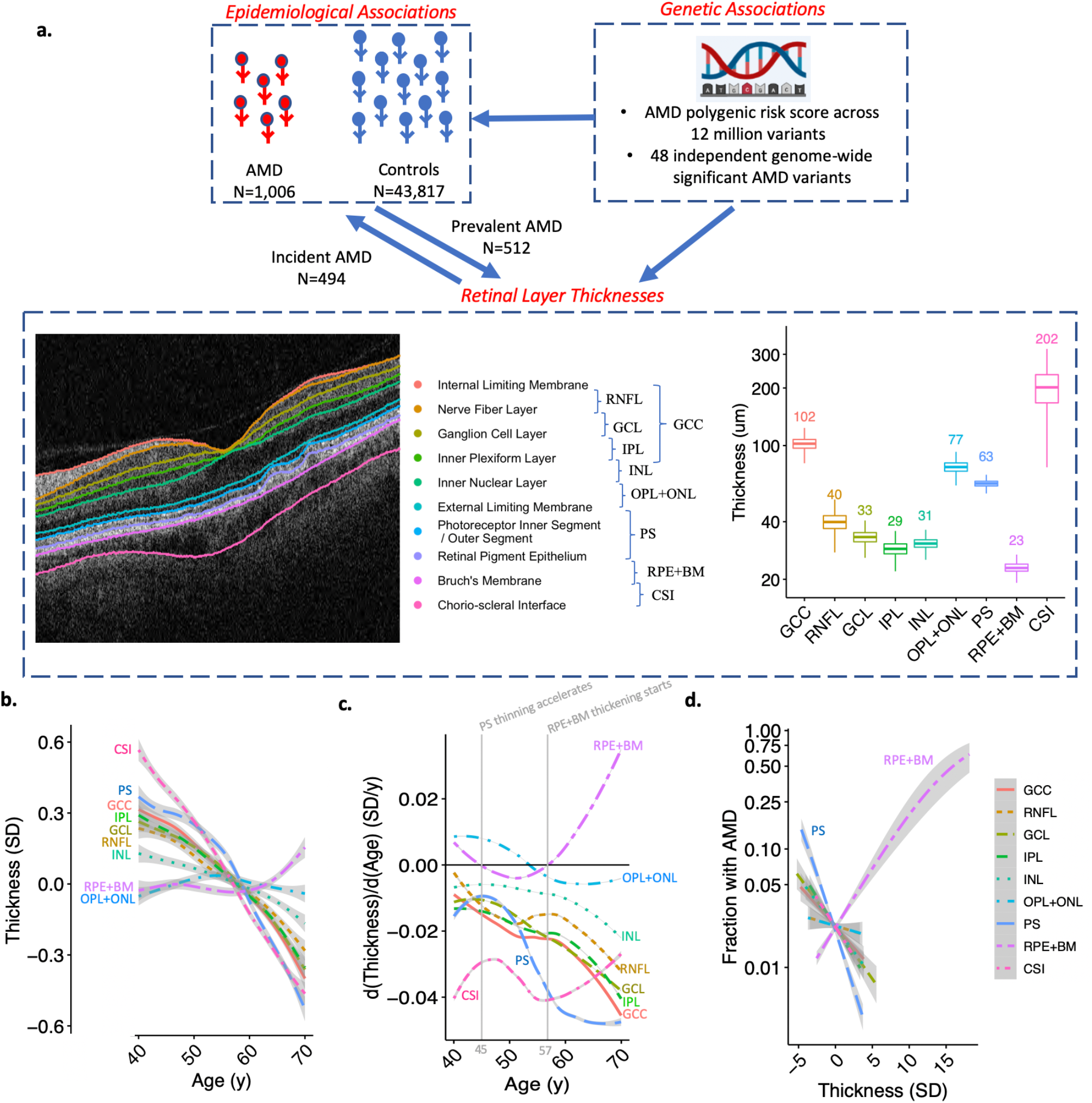
Study schema, distributions of retinal layer thicknesses and their association with age and AMD. (a) This study performed epidemiological associations of 9 retinal layer thicknesses with prevalent and incident AMD, and also genetic associations of an AMD polygenic risk score across 20 million variants, and across 48 independent genome-wide significant AMD variants from Fritsche et al. Nature Genetics 2016. Distribution of retinal layer thicknesses in micro-meters is displayed to the right of the retinal layer schematic. (b) Association of age with retinal layer thickness. Curves and standard errors reflect the best-fit generalized additive model (gam) to the individual-level data, with added smoothness. (c) Change in thickness by age for each retinal layer, plot was developed by taking the derivative (dThickness/dAge) of the plot in panel (b). Solid vertical gray line represents the age at which PS thinning accelerates (45y), and the age at which RPE+BM thickening starts (57y) across the population. (d) Association of retinal layer thickness with AMD across 9 layers. GCC = ganglion cell complex, RNFL = retinal nerve fiber layer, GCL = ganglion cell layer, IPL = inner plexiform layer, INL = inner nuclear layer, OPL+ONL = outer plexiform layer, PS = photoreceptor segment layer, RPE+BM = retinal pigment epithelium plus Bruch’s membrane, CSI = choroid scleral interface.

### Epidemiological association of retinal layer thicknesses with AMD

PS thinning and RPE+BM thickening were significantly associated with increased AMD prevalence and incidence (**Figure 1d, Figure 2**). In particular, in analyses adjusted for age, age^2^, sex, smoking status, and principal components of ancestry, each SD decrease in PS was associated with an increased odds of prevalent AMD (OR 1.37, 95% CI 1.25-1.49, P=2.5×10^−12^), and an increased risk of incident AMD (HR 1.35, 95% CI 1.23-1.47, P=3.7×10^−11^). Each SD increase in RPE+BM was associated with increased odds of prevalent AMD (OR=1.34, 95% CI 1.27-1.41, P=8.4×10^−28^) and increased risk of incident AMD (OR=1.14, 95% CI 1.06-1.22, P=0.00024). Moreover, PS thinning and RPE+BM thickening were independently significantly associated with prevalent and, separately, incident AMD when in the same model (**Supplementary Table 3**). While no associations were present between retinal layer thicknesses of other layers and prevalent AMD, there was a significant association between each SD decrease in IPL and INL layer thickness and increased risk of incident AMD (IPL: HR 1.20, 95% CI 1.10-1.32, P=0.0001; INL: HR 1.19, 95% CI: 1.09-1.30, P=0.00018) (**Figure 2c**), which was specific to males in sex-stratified analyses (IPL: HR 1.33, 95% CI 1.16-1.54; P=5.6×10^−5^; INL: HR 1.41, 95% CI 1.22-1.64, P=4.1×10^−6^) (PSex-heterogeneity <0.05) (**Supplementary Figure 2**). Further adjustment for prevalent glaucoma and myopia did not significantly change the prevalent and incident associations (**Supplementary Figures 3-6**), and no other associations with significant sex heterogeneity were identified. Sensitivity analyses were performed further adjusting for type 2 diabetes mellitus and spherical equivalent in addition to glaucoma and myopia for both prevalent and incident associations (**Supplementary Table 4**) and removing individuals with prevalent glaucoma, myopia, type 2 diabetes while also adjusting for spherical equivalent to assess the stability of the IPL and INL associations (**Supplementary Table 5**), with consistent associations observed across all analyses.

**Figure 2:**
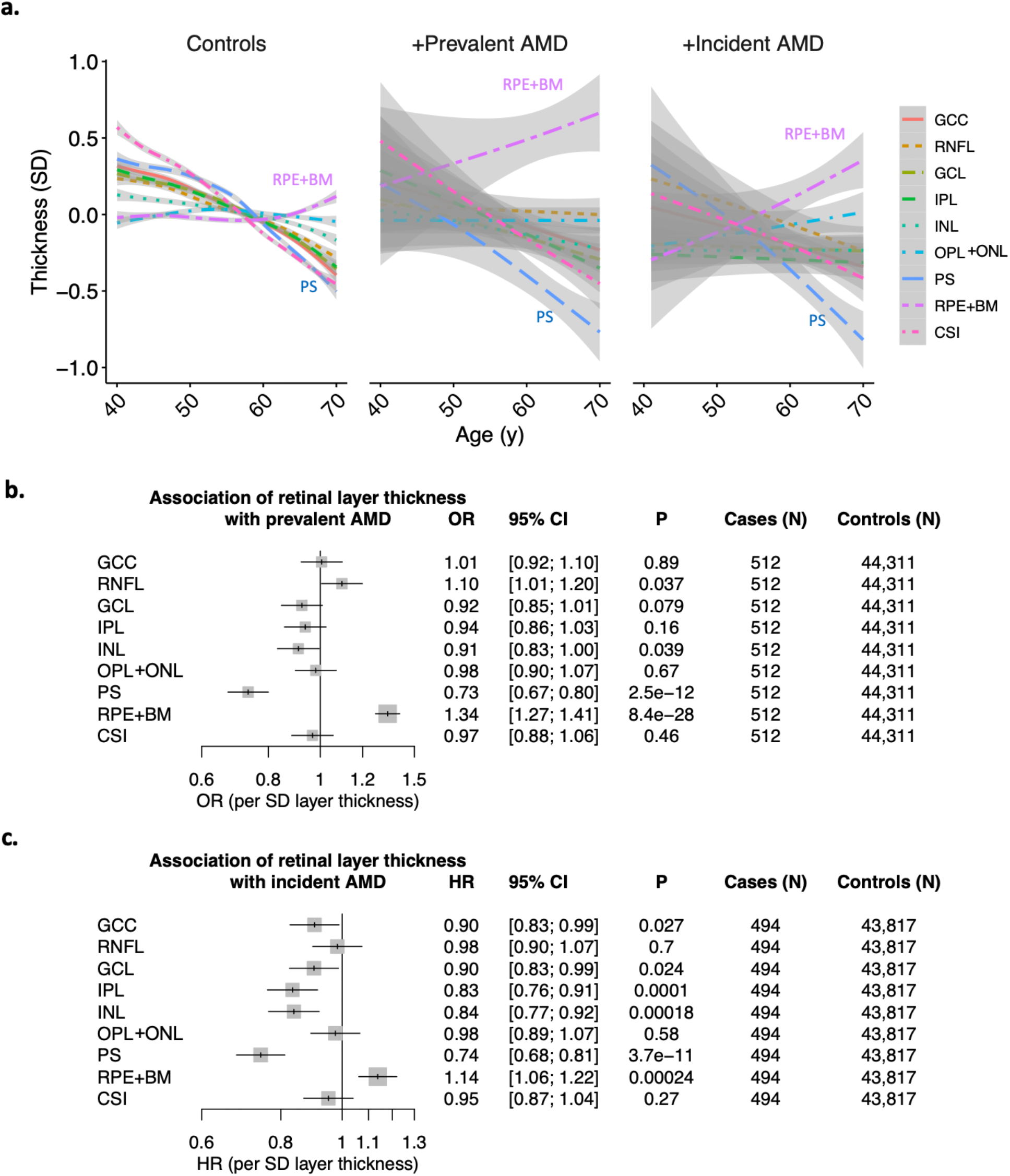
Associations of retinal layer thicknesses with prevalent and incident AMD. (a) Relationship of age with layer thickness across 9 layers of the retina, stratified by AMD status into controls, individuals with prevalent AMD at the time of OCT image acquisition, and individuals who developed incident AMD during the median 10 year follow-up. Curves and standard errors reflect the best-fit generalized additive model (gam) to the individual-level data, with added smoothness. Associations of retinal layer thickness with (b) prevalent AMD in logistic regression analyses, and (c) incident AMD in Cox proportional hazards analyses. Associations presented in panels (b) and (c) are adjusted for sex, age, age^2^, smoking status, and principal components of genetic ancestry. GCC = ganglion cell complex, RNFL = retinal nerve fiber layer, GCL = ganglion cell layer, IPL = inner plexiform layer, INL = inner nuclear layer, OPL+ONL = outer plexiform layer, PS = photoreceptor segment layer, RPE+BM = retinal pigment epithelium plus Bruch’s membrane, CSI = choroid scleral interface.

### Genetic association of an AMD polygenic risk score with retinal layer thicknesses

A genome-wide AMD polygenic risk score (PRS) was developed in the UK Biobank using summary statistics from over 12 million genetic variants analyzed in Fritsche et al. Nature Genetics 2016^14^. The scaled AMD PRS was significantly associated with AMD in the UK Biobank (across 10,751 AMD cases and 437,767 controls), with each 1 unit increase in the PRS being associated with a 2-fold increased odds of AMD (95% CI 1.90-2.10, P=2.9×10^−170^) across all individuals age 40-70 years in the UK Biobank (**Figure 3a, Supplementary Table 6**). Slightly higher effect estimates of the AMD PRS were observed among females (OR 2.11, 95% CI 1.98-2.25, P=72×10^−116^) compared to males (OR 1.86, 95% CI 1.73-2.01, P=1.3×10^−58^) (Pheterogeneity = 0.015), after adjustment for age, age^2^, smoking status, and principal components of genetic ancestry (**Supplementary Figure 7, 8**). There was a strong interaction between the AMD PRS and age (P=5.6×10^−24^) (**Supplementary Table 7**), with the AMD PRS being most strongly associated with AMD among older individuals. In particular, individuals aged 65-70 years in the 99^th^ percentile of AMD PRS were at 4.29-fold increased risk of AMD (95% CI 3.60-5.11, P=4.8×10^−59^) compared to others in the same age category (**Supplementary Figure 8b**). No significant differences in the association of the AMD PRS with AMD were observed by sex within the same 5-year age category (**Supplementary Figures 9-10**).

**Figure 3:**
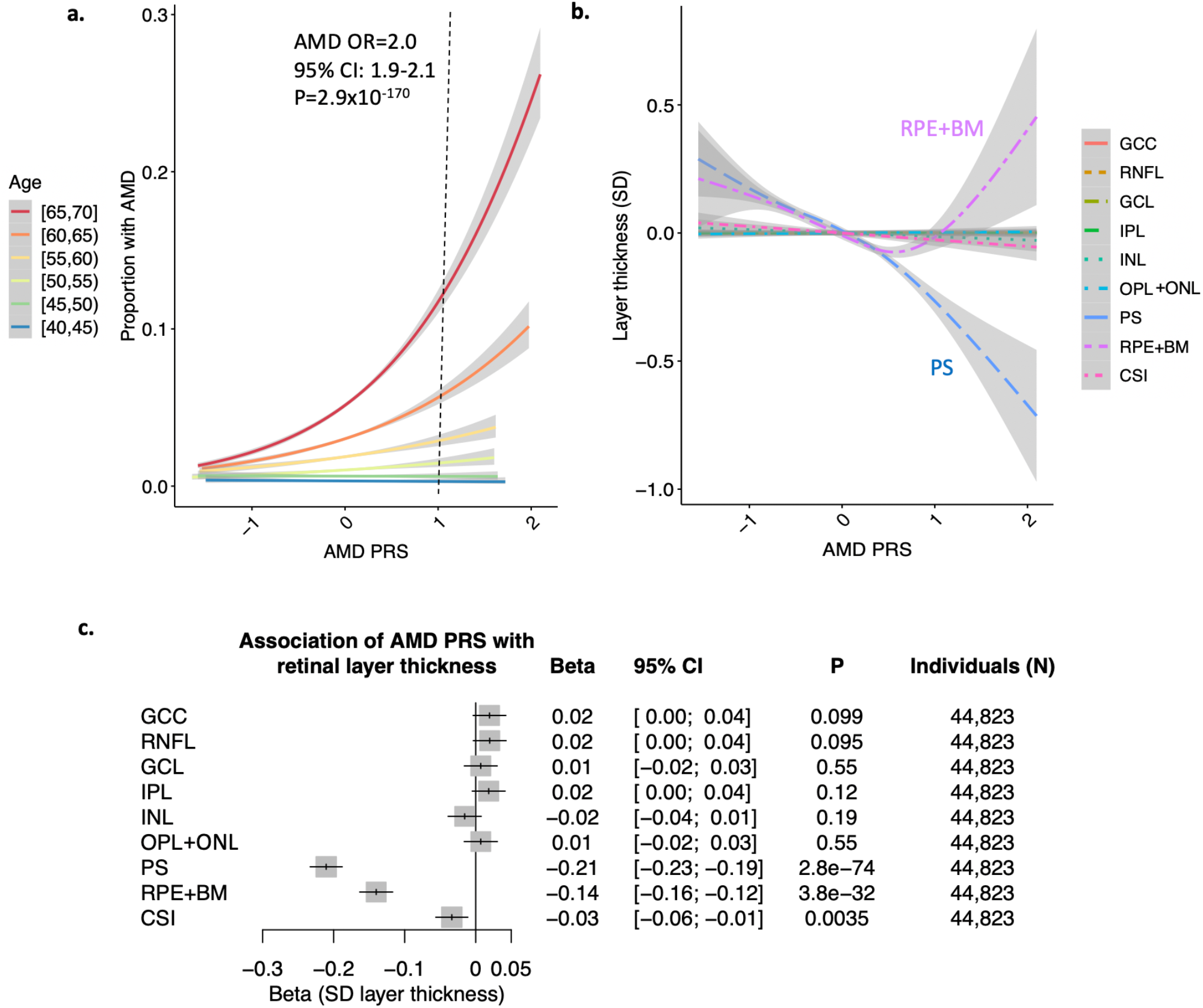
Associations of the AMD PRS with retinal layer thicknesses. (a) association of AMD PRS with AMD (prevalence + incident AMD) by 5-year age categories at enrollment. A 1 unit increase in the AMD PRS confers a 2-fold increased risk of AMD (95%CI 1.9-2.1, P=2.9×10^−170^), after adjustment for sex, age, age^2^, smoking status, and principal components of genetic ancestry. Further adjusted associations by age category are provided in **Supplementary Figure 7**. (b) relationship of AMD PRS with layer thickness highlights strong negative correlation with the PS and a U-shaped relationship with the RPE layer. Curves and standard errors in panels (a) and (b) reflect the best-fit generalized additive model (gam) to the individual-level data, with added smoothness. (c) association of AMD PRS with retinal layer thickness, adjusted for sex, age, age^2^, smoking status, and principal components of genetic ancestry. GCC = ganglion cell complex, RNFL = retinal nerve fiber layer, GCL = ganglion cell layer, IPL = inner plexiform layer, INL = inner nuclear layer, OPL = outer plexiform layer, PS = photoreceptor segment layer, RPE+BM = retinal pigment epithelium plus Bruch’s membrane layer, CSI = choroid scleral interface.

The AMD PRS was strongly associated with PS thinning and had a bimodal U-shaped relationship with RPE+BM thickness, with RPE+BM thinning with AMD PRS<0.56 (92^nd^ percentile) and thickening with AMD PRS>0.56 (**Figure 3b**). In linear regression models adjusted for age, age^2^, sex, smoking status, and principal components of ancestry, each unit increase in the AMD PRS was significantly associated with thinning of the PS (Beta -0.21 SD, 95% CI -0.23 to -0.19, P=2.8×10^−74^), RPE+BM (Beta -0.14 SD, 95% CI -0.16 to -0.12, P=3.8×10^−32^), and CSI (Beta - 0.03 SD, 95% CI -0.06 to -0.01, P=0.0035) (**Figure 3c**). Sensitivity analysis additionally removing prevalent or incident AMD cases, and adjusting for spherical equivalent in the association of the AMD PRS with retinal layer thicknesses maintained consistent associations (**Supplementary Table 8**). Further age-and sex-stratified analyses showed that females in the >99^th^ percentile of the AMD PRS who were over 60 years old had particularly thinner PS and thicker RPE+BM layers (**Supplementary Figure 11**). A focused analyses of the 35 females >60years old in the 99^th^ percentile of the AMD PRS with RPE+BM thickness >23.5um found that 22 (63%) had evidence of drusen on OCT imaging as determined by a board-certified retina specialist. More broadly, there was evidence of PS thinning (as evidenced by decreasing slope of the age association) even at age 40 for individuals among AMD PRS >20^th^ percentile, while RPE+BM thickening was not overtly apparent until age 55 among those with AMD PRS>95^th^ percentile (**Supplementary Figure 11**).

### Association of specific AMD variants with retinal layer thicknesses

To evaluate how specific loci influence retinal layer thickness, further locus-specific analyses were performed across 48 independent variants from 34 loci previously linked to advanced AMD in Fritsche et al. Nature Genetics 2016^14^, in models adjusted for age, age^2^, sex, smoking status, and principal components of genetic ancestry (**Figure 4, Supplementary Table 9**). Across the PS and RPE+BM layers, AMD risk alleles were generally associated with layer thinning (**Figure 4a**). However, different AMD risk-raising variants had heterogenous effects on retinal layers (**Figure 4b**). While there was an overall relationship between AMD risk-raising alleles and PS thinning, only AMD risk-raising variants *CFH*:rs570618-T, *CHF*:10922109-C, *CFH*:rs187328863-T, and *ARMS2*:rs3750846-C were genome-wide significantly associated with PS thinning. Interestingly, AMD risk-raising alleles *RDH5/CD63*:rs3138141-A, *PILRB/PILRA*:rs7803453-T, and *NPLOC4/TSPAN10*:rs6565597-T were conversely genome-wide significantly associated with PS thickening, raising the possibility of PS-independent AMD risks for these loci. Similarly for the RPE+BM, only the *SYN3/TIMP3*:rs5754227-T variant was genome-wide significantly associated with RPE+BM thickening; several other AMD risk variants were associated with RPE+BM thinning, including *VEGFA*:rs943080-T, *CFH*:rs570618-T, *CFH*:rs10922109-C, and *NPLOC4/TSPAN10*:rs6565597-T. Besides the PS and RPE+BM layers, several other genome-wide significant associations were also identified between AMD risk variants and thickening of the inner layers of the retina for *NPLOC4/TSPAN10*:rs6565597-T, OPL+ONL thickening for *RAD51B*:rs61985136-T, and thickening of GCL, INL, and OPL+ONL layers for *RDH5/CD63*:3138141-A. Furthermore, for CSI, the AMD risk variant *APOE*:rs429358-T was genome-wide significantly associated with CSI thinning, while the AMD risk variant *TNFRSF10A*:rs13278062-T was genome-wide significantly associated with CSI thickening (**Figure 4b, Supplementary Table 9**).

**Figure 4:**
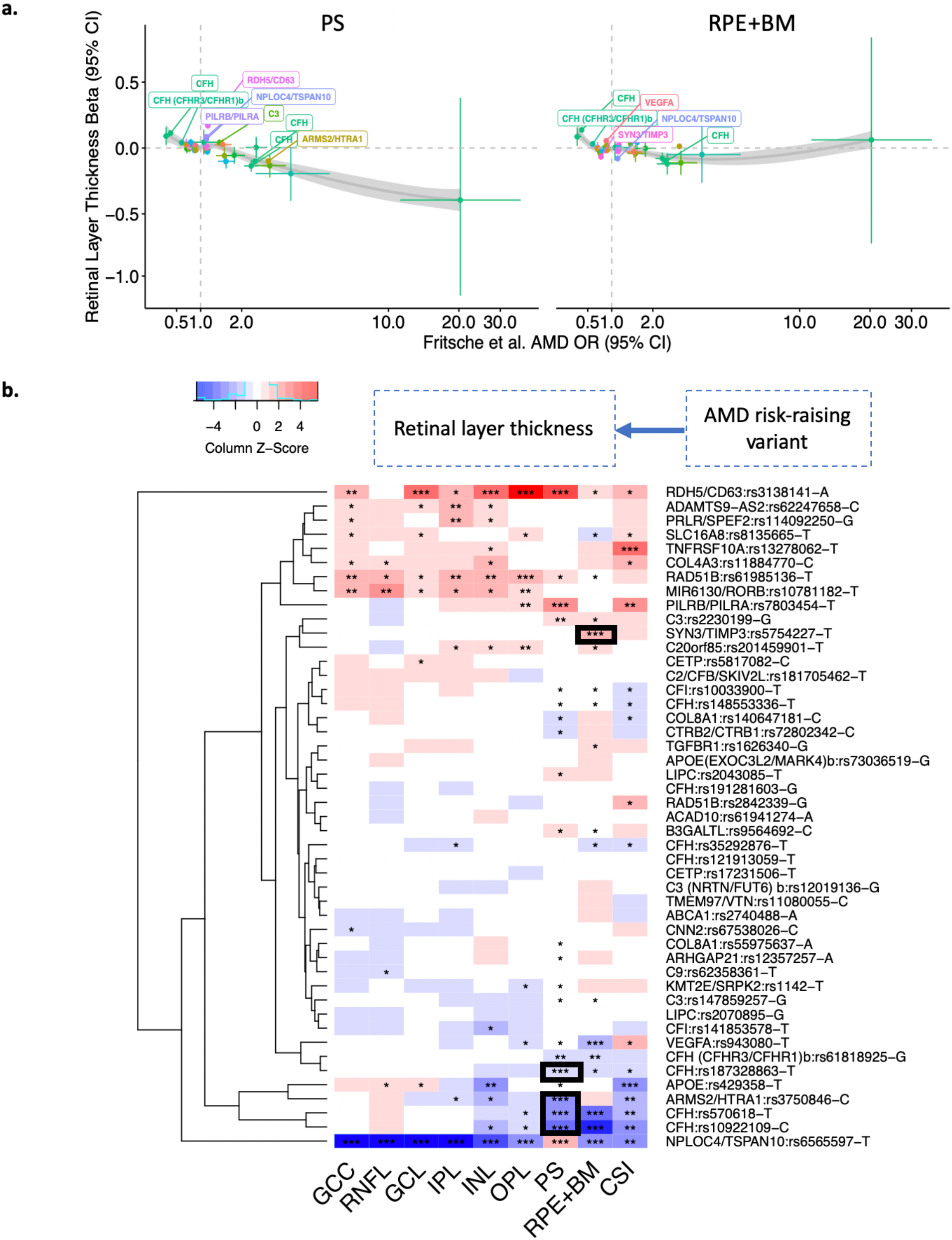
Association of AMD risk-raising variants with retinal layer thickness. (a) Plot of 48 independent variants associated with AMD (from Fritsche et al. Nature Genetics 2016) and their respective associations with PS and RPE+BM thickness in SD. Labeled genes reflect the variants with P<1×10^−4^ for layer thickness. Curves and standard errors in panel (a) are from local polynomial regression fitting. (b) association of 48 individual AMD-risk-raising variants with 9 retinal layer thicknesses. Colors reflect z-scores (beta/se) of association for each AMD-risk-raising allele with the respective retinal layer thickness. ***: P<5×10^−8^; **: 5×10^−8^<P<1×10^−4^ ; *: 1×10^−4^<P<0.05. The black boxes reflect AMD risk variants genome-wide significantly associated with PS thinning and RPE+BM thickening. A table of all associations are all provided in **Supplementary Table 9**.

### Prediction of future AMD risk improves with addition of retinal OCT-derived thicknesses

Lastly, a tiered approach to prediction of future AMD risk was assessed. A baseline model utilizing age, age^2^, sex, smoking status, body mass index, and the first ten principal components of genetic ancestry in a Cox proportional hazards model for incident AMD resulted in an area under the curve (AUC) of 0.729 (**Supplementary Figure 12**). A tiered approach successively adding on the AMD PRS (AUC 0.738), PS and RPE+BM layer thicknesses (AUC 0.748), and lastly thicknesses of all retinal layers (AUC 0.755), showed successive significant improvement in AUC with each addition (P<0.05 between each successively added variable(s)) (**Supplementary Figure 12**).

## Discussion

In summary, we jointly analyzed genomic, electronic health record, and retinal OCT data across nearly 45,000 individuals from the UK Biobank to analyze the epidemiological and genetic connections between AMD and retinal layer thicknesses (**Figure 5**). Through epidemiological analyses across 9 segmented layers of the retina, we identified trends between age and AMD that highlighted the role of photoreceptor thinning, followed by RPE+BM thickening in AMD pathogenesis. Furthermore, through genome-wide analyses, we identified a causal connection of genome-wide variants that influence AMD risk with photoreceptor segment thinning, and a heterogenous effect of AMD variants on RPE+BM thickness, with thickening of the layer specifically among individuals in the top 8 percentile of genetic risk for AMD, suggestive of drusen. Together these findings permit several conclusions.

**Figure 5:**
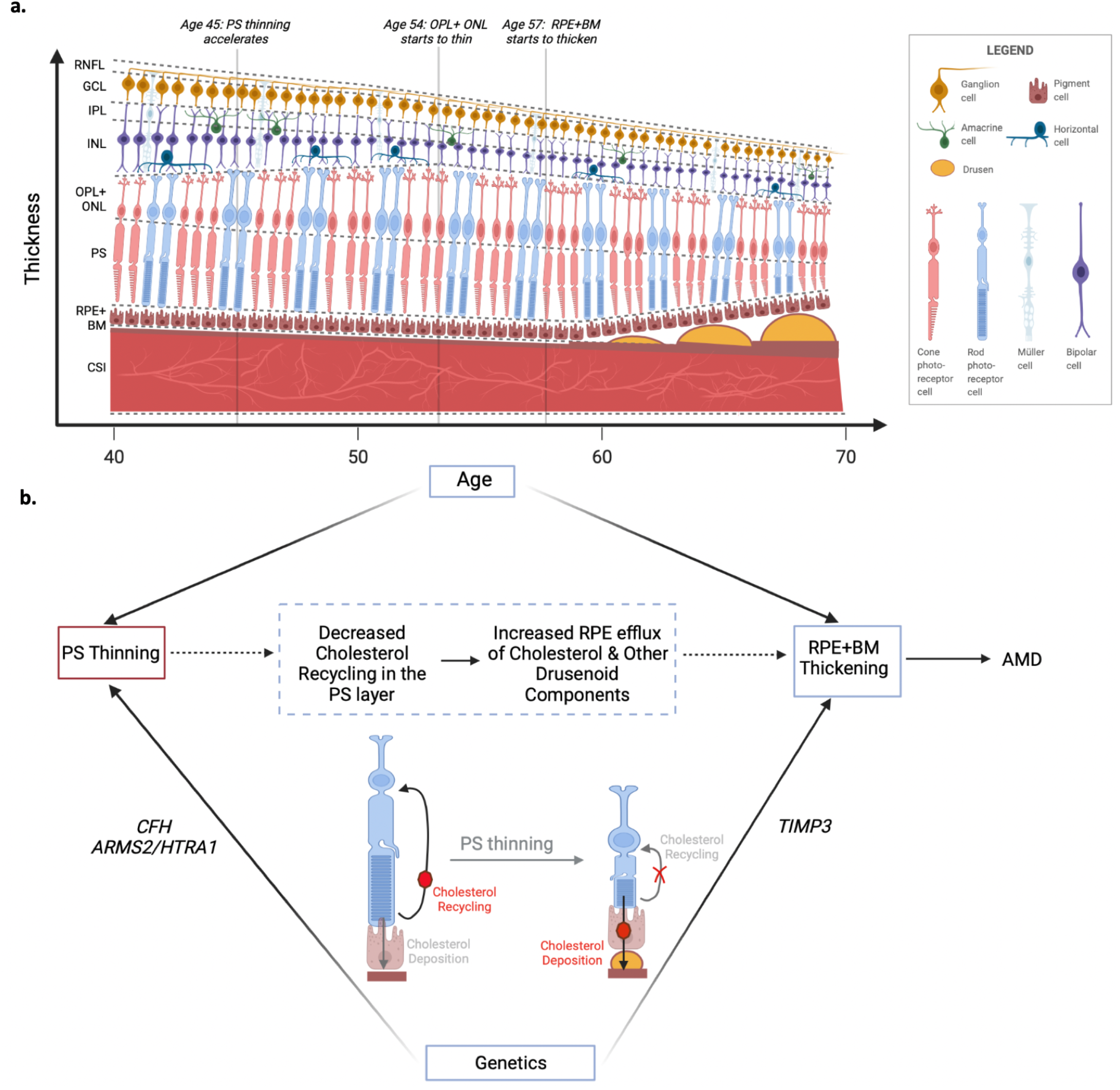
Summary schematic. a) Visualizing age-related changes in the retinal layer thicknesses. Population-wide thinning of the RNFL, GCL, IPL, INL, and CSI occurs across ages 40-70, PS thinning accelerates at age 45, OPL+ONL thinning starts at age 54, and RPE+BM thickening starts at age 57. While RPE+BM thickening may relate in part to drusen development, it may also reflect basal laminar deposits in the BM, or other generalized BM thickening, and/or RPE thickening. b) PS thinning precedes RPE+BM thickening by decades. Both PS thinning and RPE+BM thickening are epidemiologically associated with prevalent and incident AMD. Additionally, several inherited AMD genetic risk loci also are associated with PS thinning (at the *CFH* and *ARMS2/HTRA1* loci), and with RPE+BM thickening (at the *TIMP3* locus). One mechanistic hypothesis linking PS thinning with RPE+BM thickening is through decreased cholesterol recycling in the PS outer segment leading to increased cholesterol deposition into the Bruch’s membrane which over time develops into drusen deposits. Figure created using BioRender.com. RNFL = retinal nerve fiber layer, GCL = ganglion cell layer, IPL = inner plexiform layer, INL = inner nuclear layer, OPL+ONL = outer plexiform layer, PS = photoreceptor segment layer, RPE+BM = retinal pigment epithelium plus Bruch’s membrane, CSI = choroid scleral interface.

First, epidemiological analyses of population-wide trends of retinal layer thickness with age and AMD status suggests PS thinning may precede RPE+BM thickening by decades, and that thicknesses of both layers are useful in predicting future risk of AMD. While prior work had similarly linked drusen development in the RPE+BM layer and photoreceptor degeneration with AMD, it has thus far been unclear which occurs first. Some studies hypothesized that photoreceptors, which aid in cholesterol turnover in the outer retina, are key to recycling cholesterol, and impaired recycling (through mechanisms including photoreceptor degeneration or thinning) could increase cholesterol load onto the RPE and thereby increase lipid and protein excretion from the basal RPE contributing towards drusen development^25^ (**Figure 5b**). In the reverse direction, other studies have hypothesized that hypoxia in the photoreceptor layer, secondary to drusen development, may lead to photoreceptor degeneration^26^. Here, we clearly observe thinning of the PS layer across all ages of individuals in this study (40-70y), while thickening of the RPE+BM layer on average progressed after age 57 years, suggesting that PS thinning precedes RPE+BM thickening among the general population (**Figure 1b,c**), though certainly thickening of the RPE+BM also plays a role in AMD disease progression. Thickening of the RPE+BM layer was significantly associated with a 34% increased odds of prevalent AMD and 14% increased risk of future incident AMD diagnosis. This finding is consistent with the notion that this layer is enlarged by drusen and BLamD in AMD, although other possible mechanisms for RPE+BM thickening also include diffuse thickening of the BM or RPE. Additionally, each SD decrease in PS thickness was significantly associated with 37% increased odds of prevalent AMD and 35% increased risk of future incident AMD diagnosis. Indeed, prior reports have also observed photoreceptor loss in both early and advanced AMD^6,7^. These results together indicate that PS and RPE+BM are biomarkers of future AMD risk, with PS thinning preceding RPE+BM thickening.

Second, epidemiological analyses also identified possible contributors of IPL and INL thinning to increased risk of future incident AMD specifically in males, independent of myopia, glaucoma, type 2 diabetes mellitus, and spherical equivalent. This may suggest a possible sex-specific component to AMD in males related to cells in these layers, namely bipolar cells and other supportive interneurons (amacrine, horizontal cells) restricted to the IPL/INL layers that synapse with photoreceptors basally and ganglion cells apically, or Muller cells that traverse the IPL/INL layers. A prior study (N=90), similarly found that thinning of the IPL layer was associated with AMD^11^. However, this finding was not replicated in a larger study (N= 449)^8^. Here we show evidence across a large population cohort of IPL and INL thinning preceding AMD diagnosis in males. Further work needs to be performed to understand the sex-specific nature of the association of IPL/INL thinning with incident AMD in males.

Third, while several small studies (N<100) have suggested that GCC and RNFL layer thinning are also associated with AMD^9,10^, no association between GCC and RNFL layer thickness with prevalent or incident AMD was detected in this study, suggesting that the association from the smaller studies may have been false positives or confounded by other factors (including segmentation quality^11^). Alternatively, it is possible that the imaging modality, layer segmentation and analysis methods, and/or population utilized in prior studies that identified associations between GCC thinning and AMD were significantly different from ours. In either case, our finding is consistent with a histopathologic study that found that the number of ganglion cells was similar in 76 eyes with dry AMD versus 76 controls^27^.

Fourth, genetic analyses identified a significant association between genetically increased risk for AMD and photoreceptor thinning, while a bimodal association was detected with RPE+BM thickness, suggestive of drusen development with AMD PRS over the 90^th^ percentile. In particular, locus-specific analyses identified potential mechanistic roles of AMD risk alleles at *CFH* and *ARMS2/HTRA1* on PS thinning. Prior work has suggested that while variants in *CFH* and *ARMS2/HTRA1* loci were not associated with macular thickness averaged within the outermost circle of the ETDRS map among UK Biobank participants^15^, these loci are associated with peri-foveal changes in macular retinal thickness on OCT^16^ (N=299). However, these studies did not examine specific retinal layers individually. In-vivo studies have shown that CFH -/- knockout mice express complement C3 in the outer segments, while control mice only have C3 detectable at the BM^28^. Furthermore, CFH -/- knockout mice also consistently had misaligned, disorganized photoreceptors with bent and sometimes horizontal outer segments, and with loss of the close proximity between normally perpendicular PS outer tips and apical microvilli of the RPE^28^. Similar PS changes have been reported in eyes from elderly humans^29^ reflecting an imbalance between PS outer segment formation and outer segment disc turnover and phagocytosis. As for the *ARMS2/HTRA1* locus association, *ARMS2* is expressed in human RPE cells (ARPE-19), and in-vitro experiments suggest that the *ARMS2* plays a role in the phagocytosis of PS outer segments in RPE cells^30^. The expression of the alternative gene at this locus, *HTRA1*, in both PS and in the RPE, increases with aging and with AMD disease-associated variants in zebrafish models^31^. PS-specific overexpression of HTRA1 in zebrafish resulted in caspase-induced cell death of photoreceptors, which was rescued by the suppression of htra1 by the inhibitor 6-boroV^31^. The genome-wide association of AMD genetic risk with photoreceptor thinning suggests that photoreceptor thinning may be a causal mechanism through which these genetic loci increase risk of AMD. However, we acknowledge that the relationship between AMD and PS thinning may be bidirectional, in the sense that independent genome-wide significant variants that influence AMD risk are associated with PS thinning, and also that independent genome-wide significant variants that influence PS thinning may similarly increase risk of AMD. Further studies are needed to elucidate this.

Locus-specific associations for AMD loci also identified AMD-risk raising alleles at *SYN3/TIMP3* and their association with RPE+BM thickening, indicative of drusen development. Prior work has suggested the *TIMP3* gene, which encodes the tissue inhibitor of metalloproteinase 3 protein, as the likely causal gene given its high expression in human fetal RPE cells and the significant association of the respective protective AMD GWAS variant (rs9621532-C) with decreased transcription of TIMP3 mRNA in human fetal RPE cells^32^. Functionally, TIMPs inhibit matrix metalloproteinases, peptides that degrade extracellular matrix. *TIMP3* mutations have been implicated in Sorsby’s fundus dystrophy, an autosomal dominant disorder featuring accumulation of macular drusen and progression to neovascularization and retinal degeneration^33^. Additionally, TIMP3 protein expression in the Bruch’s membrane increases with age^34^, and TIMP3 protein levels in Bruch’s membrane and in drusen are correlated with AMD disease pathology^35^. Thus, *TIMP3* could contribute towards thickening of the Bruch’s membrane and drusen development secondary to impaired clearance of basally-excreted RPE by-products through the Bruch’s membrane^35^.

Fifth, published AMD risk calculators^36^ have relied on conventional demographic data (age, sex, smoking status, BMI, race), and select AMD genetic markers (https://www.seddonamdriskscore.org/amd/amd.jsp?action=NEW). Here, we show that the inclusion of layer thickness data in addition to demographic phenotypes (age, sex, smoking status, BMI, and genetic ancestry), and a genome-wide polygenic risk score significantly improves prediction of incident AMD. Furthermore, the inclusion of data across all retinal layers significantly improves incident AMD prediction, compared to inclusion of data from only the PS and RPE+BM layers, suggesting that the addition of other layers (such as the aforementioned IPL/INL layers) significantly adds to an incident AMD prediction model.

Although our study has several strengths, there are important limitations to consider. First, while our analyses were performed adjusted for age, age^2^, sex, smoking status, and genetic ancestry, axial length was not available for adjustment of the OCT retinal layer associations; however sensitivity analyses additionally adjusting for spherical equivalent measurements showed consistent associations. For associations of INL/IPL with incident AMD, additional removal of individuals with myopia, glaucoma, and type 2 diabetes mellitus diagnoses at enrollment was performed, in addition to adjustment for spherical equivalent, which also did not mitigate the respective associations. Second, due to genetic analyses performed, the present analyses used Europeans in the UK Biobank; further analyses in diverse ethnic cohorts and would enable a more comprehensive assessment of the associations across ethnicities. Third, while our definition of prevalent and incident AMD combined all available information from the UK Biobank, including ICD-9, ICD-10, self-report, and general practice data, this definition suffers from inherent imprecision; however the size of our dataset likely partially mitigates this problem. Fourth, it is possible that contributors to image quality including media opacities may influence and confound the phenotypic and genotypic associations. Robust and conservative efforts were taken towards exclusion of poor-quality OCT segmentations. Though it remains possible that poor-quality segmentations remain in the dataset, the large size of our dataset and robustness of associations between outer layer segment thicknesses and AMD make it unlikely to contribute towards false-positive associations.

In conclusion, our epidemiological analyses suggest that photoreceptor thinning may precede RPE+BM thickening in AMD by decades, and that PS thinning is the strongest retinal layer in predicting future risk of AMD. Additionally, genetic variants that increase risk of AMD are significantly associated with decreased PS thickness, while a bimodal association was detected with RPE+BM thickness, suggestive of drusen and BLamD development with AMD PRS over the 90^th^ percentile. Furthermore, locus-specific analyses identified potential mechanistic roles of AMD risk alleles at *CFH* and *ARMS2/HTRA1* on PS thinning and at *SYN3/TIMP3* on RPE+BM thickening. Overall, these findings support a potential causal influence of AMD genetic variants on PS thinning, and suggest the clinical use of PS thinning as an early-stage biomarker for risk of future AMD development.

## Data Availability

UKB individual-level data are available for request by application (https://www.ukbiobank.ac.uk).

