## Supplementary Material for "Photoreceptor layer thinning is an early biomarker for age-related macular degeneration development: Epidemiological and genetic evidence from UK Biobank optical coherence tomography data"

Supplementary Figures

### Table of Contents:

Supplementary Figures:


1) Supplementary Figure 1: Association of age with retinal layer thickness.
2) Supplementary Figure 2: Heterogeneity of the association of retinal layer thicknesses with incident AMD by sex.
3) Supplementary Figure 3: Heterogeneity of the association of retinal layer thicknesses with incident AMD by sex, additionally adjusting for prevalent glaucoma and myopia.
4) Supplementary Figure 4: Heterogeneity of the association of retinal layer thicknesses with prevalent AMD by sex.
5) Supplementary Figure 5: Heterogeneity of the association of retinal layer thicknesses with prevalent AMD by sex, additionally adjusting for prevalent glaucoma and myopia.
6) Supplementary Figure 6: Association of retinal layer thicknesses with prevalent and incident AMD, additionally adjusting for prevalent glaucoma and myopia.
7) Supplementary Figure 7: Association of AMD PRS with AMD stratified by age.
8) Supplementary Figure 8: Association of AMD PRS with AMD stratified by sex and age.
9) Supplementary Figure 9: Association of AMD PRS with AMD in (a) Females and (b) Males, by 5-year age groups.
10) Supplementary Figure 10: Heterogeneity of association of the AMD PRS with AMD by sex among different age groups.
11) Supplementary Figure 11: Relationship of age with (a) photoreceptor and (b) RPE+BM layer thicknesses by sex and AMD PRS percentile.
12) Supplementary Figure 12: Prediction of incident AMD significantly improves with addition of retinal layer thicknesses from OCT, on top of baseline characteristics and a polygenetic risk score.

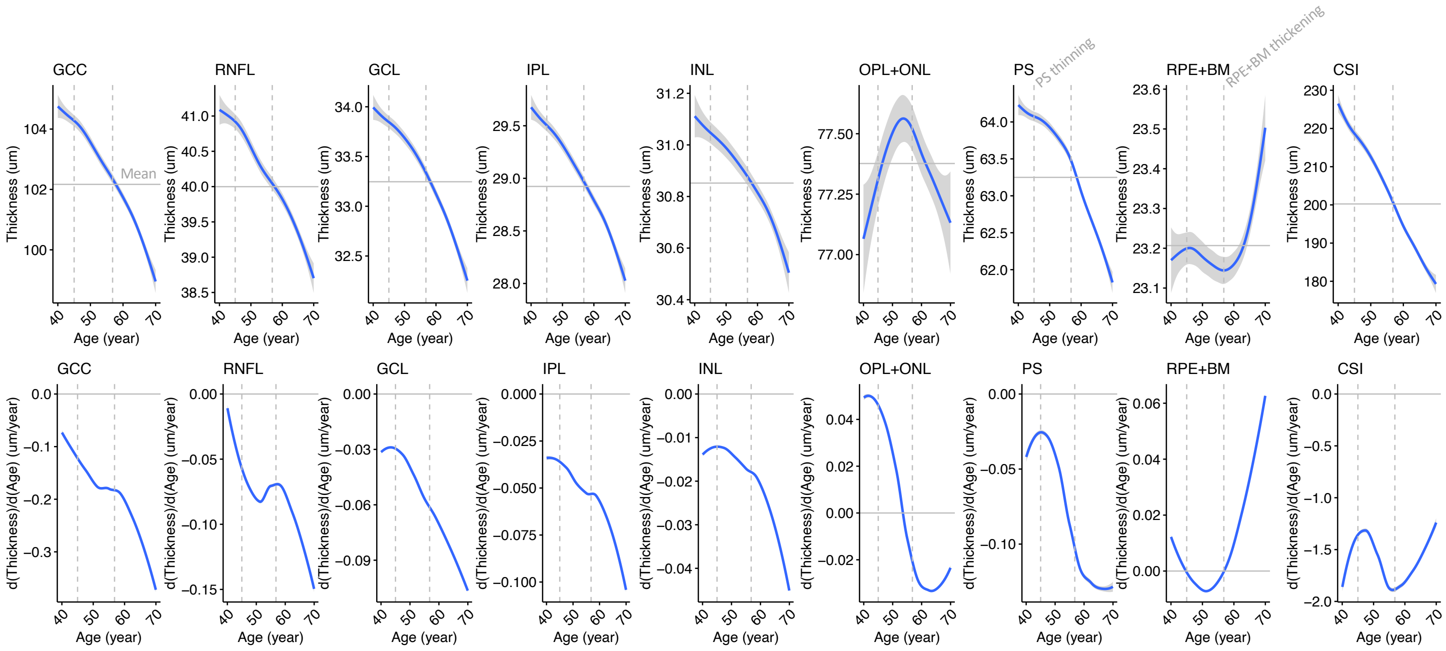


Supplementary Figure 1: Association of age with retinal layer thickness. (a) Relationship of age with retinal layer thickness. Curves and standard errors reflect the best-fit generalized additive model (gam) to the individual-level data, with added smoothness. Horizontal lines reflect the mean thickness across all studied ages. (b) Change in thickness by age for each retinal layer, plot was developed by taking the derivative (dThickness/dAge) of the plot in panel (b). Solid vertical gray line represents the age at which PS thinning accelerates (45y), and the age at which RPE+BM thickening starts (57y) across the population. GCC = ganglion cell complex, RNFL = retinal nerve fiber layer, GCL = ganglion cell layer, IPL = inner plexiform layer, INL = inner nuclear layer, OPL+ONL = outer plexiform layer, PS = photoreceptor segment layer, RPE+BM = retinal pigment epithelium plus Bruch's membrane, CSI = choroid scleral interface.

| Association of retinal layer thickness with incident AMD | HR | 95% CI | P | Cases (N) | Controls (N) |
|---|---|---|---|---|---|
| **GCC** | | | | | |
| Female | 0.94 | [0.84; 1.06] | 0.32 | 286 | 23,478 |
| Male | 0.85 | [0.75; 0.98] | 0.024 | 208 | 20,339 |
| Heterogeneity: *p* = 0.28 | | | | | |
| **RNFL** | | | | | |
| Female | 0.96 | [0.85; 1.08] | 0.47 | 286 | 23,478 |
| Male | 1.01 | [0.89; 1.16] | 0.83 | 208 | 20,339 |
| Heterogeneity: *p* = 0.52 | | | | | |
| **GCL** | | | | | |
| Female | 0.97 | [0.87; 1.09] | 0.66 | 286 | 23,478 |
| Male | 0.81 | [0.70; 0.93] | 0.0027 | 208 | 20,339 |
| Heterogeneity: *p* = 0.04 | | | | | |
| **IPL** | | | | | |
| Female | 0.92 | [0.82; 1.04] | 0.19 | 286 | 23,478 |
| Male | 0.72 | [0.63; 0.84] | 8.8e−06 | 208 | 20,339 |
| Heterogeneity: *p* = 0.01 | | | | | |
| **INL** | | | | | |
| Female | 0.95 | [0.84; 1.06] | 0.35 | 286 | 23,478 |
| Male | 0.71 | [0.62; 0.82] | 4.4e−06 | 208 | 20,339 |
| Heterogeneity: *p* < 0.01 | | | | | |
| **OPL+ONL** | | | | | |
| Female | 1.01 | [0.90; 1.13] | 0.92 | 286 | 23,478 |
| Male | 0.94 | [0.82; 1.07] | 0.34 | 208 | 20,339 |
| Heterogeneity: *p* = 0.43 | | | | | |
| **PS** | | | | | |
| Female | 0.74 | [0.66; 0.83] | 3e−07 | 286 | 23,478 |
| Male | 0.75 | [0.65; 0.86] | 3.1e−05 | 208 | 20,339 |
| Heterogeneity: *p* = 0.92 | | | | | |
| **RPE+BM** | | | | | |
| Female | 1.10 | [1.00; 1.21] | 0.055 | 286 | 23,478 |
| Male | 1.21 | [1.08; 1.34] | 0.00061 | 208 | 20,339 |
| Heterogeneity: *p* = 0.19 | | | | | |
| **CSI** | | | | | |
| Female | 0.97 | [0.87; 1.09] | 0.63 | 286 | 23,478 |
| Male | 0.92 | [0.80; 1.06] | 0.27 | 208 | 20,339 |
| Heterogeneity: *p* = 0.58 | | | | | |

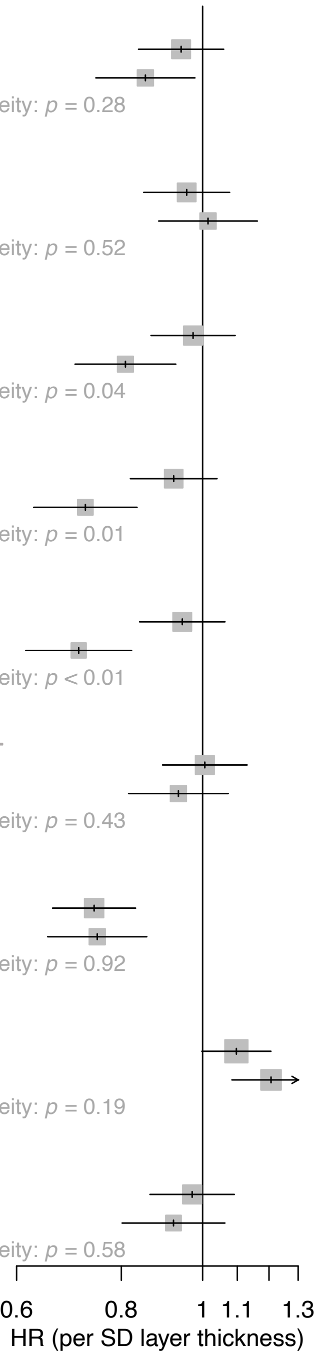


Supplementary Figure 2: Heterogeneity of the association of retinal layer thicknesses with incident AMD by sex. GCC = ganglion cell complex, RNFL = retinal nerve fiber layer, GCL = ganglion cell layer, IPL = inner plexiform layer, INL = inner nuclear layer, OPL+ONL = outer plexiform layer plus outer nuclear layer, PS = photoreceptor segment layer, RPE+BM = retinal pigment epithelium plus Bruch's membrane, CSI = choroid scleral interface.

### +Glaucoma, Myopia adjustment:

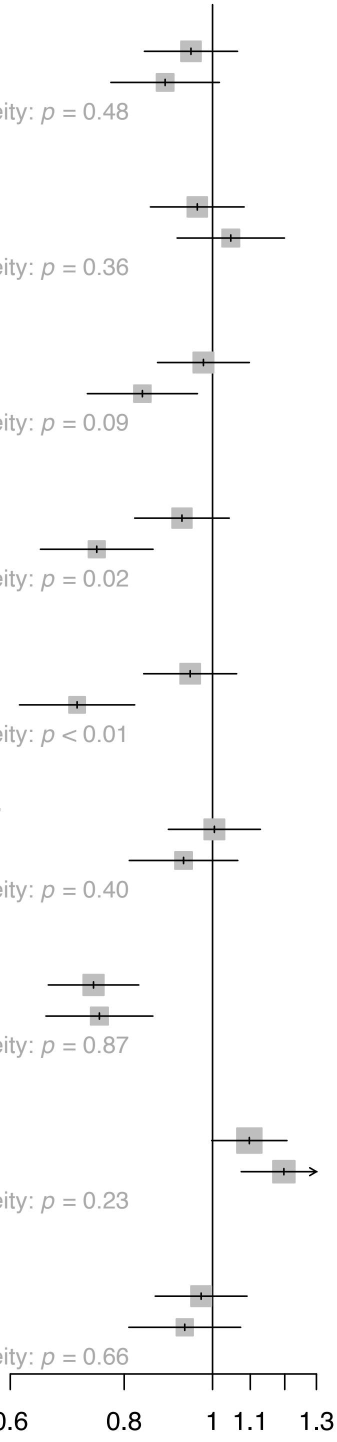


| Association of retinal layer thickness with incident AMD | HR | 95% CI | P | Cases (N) | Controls (N) |
|---|---|---|---|---|---|
| **GCC** | | | | | |
| Female | 0.95 | [0.84; 1.07] | 0.36 | 286 | 23,478 |
| Male | 0.89 | [0.77; 1.02] | 0.087 | 208 | 20,339 |
| Heterogeneity: *p* = 0.48 | | | | | |
| **RNFL** | | | | | |
| Female | 0.96 | [0.85; 1.08] | 0.52 | 286 | 23,478 |
| Male | 1.05 | [0.91; 1.20] | 0.51 | 208 | 20,339 |
| Heterogeneity: *p* = 0.36 | | | | | |
| **GCL** | | | | | |
| Female | 0.98 | [0.87; 1.10] | 0.7 | 286 | 23,478 |
| Male | 0.84 | [0.73; 0.96] | 0.012 | 208 | 20,339 |
| Heterogeneity: *p* = 0.09 | | | | | |
| **IPL** | | | | | |
| Female | 0.93 | [0.82; 1.04] | 0.2 | 286 | 23,478 |
| Male | 0.75 | [0.65; 0.86] | 5.6e−05 | 208 | 20,339 |
| Heterogeneity: *p* = 0.02 | | | | | |
| **INL** | | | | | |
| Female | 0.95 | [0.84; 1.06] | 0.35 | 286 | 23,478 |
| Male | 0.71 | [0.61; 0.82] | 4.1e−06 | 208 | 20,339 |
| Heterogeneity: *p* < 0.01 | | | | | |
| **OPL+ONL** | | | | | |
| Female | 1.00 | [0.89; 1.13] | 0.94 | 286 | 23,478 |
| Male | 0.93 | [0.81; 1.07] | 0.29 | 208 | 20,339 |
| Heterogeneity: *p* = 0.40 | | | | | |
| **PS** | | | | | |
| Female | 0.74 | [0.66; 0.83] | 2.5e−07 | 286 | 23,478 |
| Male | 0.75 | [0.66; 0.86] | 3.4e−05 | 208 | 20,339 |
| Heterogeneity: *p* = 0.87 | | | | | |
| **RPE+BM** | | | | | |
| Female | 1.10 | [1.00; 1.21] | 0.055 | 286 | 23,478 |
| Male | 1.20 | [1.08; 1.33] | 0.001 | 208 | 20,339 |
| Heterogeneity: *p* = 0.23 | | | | | |
| **CSI** | | | | | |
| Female | 0.97 | [0.87; 1.09] | 0.63 | 286 | 23,478 |
| Male | 0.93 | [0.81; 1.07] | 0.33 | 208 | 20,339 |
| Heterogeneity: *p* = 0.66 | | | | | |

Supplementary Figure 3: Heterogeneity of the association of retinal layer thicknesses with incident AMD by sex, additionally adjusting for prevalent glaucoma and myopia. GCC = ganglion cell complex, RNFL = retinal nerve fiber layer, GCL = ganglion cell layer, IPL = inner plexiform layer, INL = inner nuclear layer, OPL+ONL = outer plexiform layer plus outer nuclear layer, PS = photoreceptor segment layer, RPE+BM = retinal pigment epithelium plus Bruch’s membrane, CSI = choroid scleral interface.

**Association of retinal layer thickness with prevalent AMD**

| | OR | 95% CI | P | Cases (N) | Controls (N) |
|---|---|---|---|---|---|
| **GCC** | | | | | |
| Female | 1.03 | [0.91; 1.15] | 0.66 | 294 | 23,764 |
| Male | 0.98 | [0.86; 1.12] | 0.79 | 218 | 20,547 |
| Heterogeneity: $p = 0.63$ | | | | | |
| **RNFL** | | | | | |
| Female | 1.14 | [1.01; 1.28] | 0.03 | 294 | 23,764 |
| Male | 1.05 | [0.92; 1.20] | 0.47 | 218 | 20,547 |
| Heterogeneity: $p = 0.38$ | | | | | |
| **GCL** | | | | | |
| Female | 0.93 | [0.83; 1.04] | 0.22 | 294 | 23,764 |
| Male | 0.91 | [0.80; 1.05] | 0.19 | 218 | 20,547 |
| Heterogeneity: $p = 0.85$ | | | | | |
| **IPL** | | | | | |
| Female | 0.93 | [0.83; 1.05] | 0.23 | 294 | 23,764 |
| Male | 0.95 | [0.83; 1.09] | 0.45 | 218 | 20,547 |
| Heterogeneity: $p = 0.83$ | | | | | |
| **INL** | | | | | |
| Female | 0.89 | [0.79; 1.00] | 0.044 | 294 | 23,764 |
| Male | 0.94 | [0.82; 1.08] | 0.4 | 218 | 20,547 |
| Heterogeneity: $p = 0.50$ | | | | | |
| **OPL+ONL** | | | | | |
| Female | 0.99 | [0.88; 1.11] | 0.83 | 294 | 23,764 |
| Male | 0.97 | [0.85; 1.11] | 0.63 | 218 | 20,547 |
| Heterogeneity: $p = 0.82$ | | | | | |
| **PS** | | | | | |
| Female | 0.75 | [0.67; 0.85] | 1.3e−06 | 294 | 23,764 |
| Male | 0.70 | [0.62; 0.80] | 2.6e−07 | 218 | 20,547 |
| Heterogeneity: $p = 0.45$ | | | | | |
| **RPE+BM** | | | | | |
| Female | 1.38 | [1.30; 1.47] | 3.2e−24 | 294 | 23,764 |
| Male | 1.24 | [1.13; 1.37] | 1.7e−05 | 218 | 20,547 |
| Heterogeneity: $p = 0.08$ | | | | | |
| **CSI** | | | | | |
| Female | 0.97 | [0.87; 1.09] | 0.66 | 294 | 23,764 |
| Male | 0.96 | [0.83; 1.10] | 0.54 | 218 | 20,547 |
| Heterogeneity: $p = 0.86$ | | | | | |

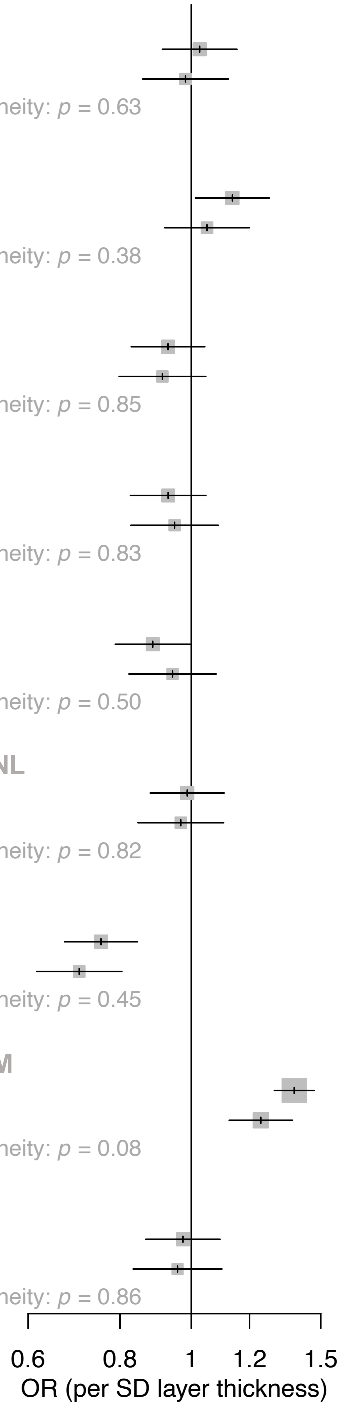


Supplementary Figure 4: Heterogeneity of the association of retinal layer thicknesses with prevalent AMD by sex. GCC = ganglion cell complex, RNFL = retinal nerve fiber layer, GCL = ganglion cell layer, IPL = inner plexiform layer, INL = inner nuclear layer, OPL+ONL = outer plexiform layer plus outer nuclear layer, PS = photoreceptor segment layer, RPE+BM = retinal pigment epithelium plus Bruch's membrane, CSI = choroid scleral interface.

### +Glaucoma, Myopia adjustment:

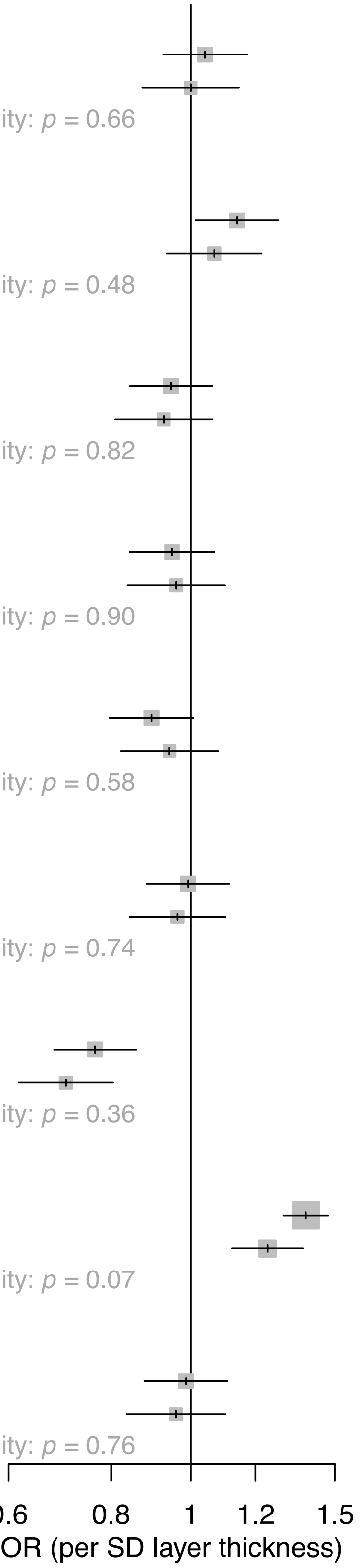


| Association of retinal layer thickness with prevalent AMD | OR | 95% CI | P | Cases (N) | Controls (N) |
|---|---|---|---|---|---|
| **GCC** | | | | | |
| Female | 1.04 | [0.93; 1.17] | 0.5 | 294 | 23,764 |
| Male | 1.00 | [0.87; 1.14] | 1 | 218 | 20,547 |
| Heterogeneity: *p* = 0.66 | | | | | |
| **RNFL** | | | | | |
| Female | 1.14 | [1.01; 1.28] | 0.028 | 294 | 23,764 |
| Male | 1.07 | [0.94; 1.22] | 0.33 | 218 | 20,547 |
| Heterogeneity: *p* = 0.48 | | | | | |
| **GCL** | | | | | |
| Female | 0.95 | [0.84; 1.06] | 0.35 | 294 | 23,764 |
| Male | 0.93 | [0.81; 1.06] | 0.28 | 218 | 20,547 |
| Heterogeneity: *p* = 0.82 | | | | | |
| **IPL** | | | | | |
| Female | 0.95 | [0.84; 1.07] | 0.39 | 294 | 23,764 |
| Male | 0.96 | [0.84; 1.10] | 0.56 | 218 | 20,547 |
| Heterogeneity: *p* = 0.90 | | | | | |
| **INL** | | | | | |
| Female | 0.90 | [0.80; 1.01] | 0.068 | 294 | 23,764 |
| Male | 0.94 | [0.82; 1.08] | 0.39 | 218 | 20,547 |
| Heterogeneity: *p* = 0.58 | | | | | |
| **OPL+ONL** | | | | | |
| Female | 0.99 | [0.88; 1.11] | 0.9 | 294 | 23,764 |
| Male | 0.96 | [0.84; 1.10] | 0.59 | 218 | 20,547 |
| Heterogeneity: *p* = 0.74 | | | | | |
| **PS** | | | | | |
| Female | 0.76 | [0.68; 0.86] | 4.9e−06 | 294 | 23,764 |
| Male | 0.70 | [0.62; 0.81] | 2.6e−07 | 218 | 20,547 |
| Heterogeneity: *p* = 0.36 | | | | | |
| **RPE+BM** | | | | | |
| Female | 1.38 | [1.30; 1.47] | 3e−24 | 294 | 23,764 |
| Male | 1.24 | [1.12; 1.37] | 2.1e−05 | 218 | 20,547 |
| Heterogeneity: *p* = 0.07 | | | | | |
| **CSI** | | | | | |
| Female | 0.99 | [0.88; 1.11] | 0.83 | 294 | 23,764 |
| Male | 0.96 | [0.84; 1.10] | 0.56 | 218 | 20,547 |
| Heterogeneity: *p* = 0.76 | | | | | |

Supplementary Figure 5: Heterogeneity of the association of retinal layer thicknesses with prevalent AMD by sex, additionally adjusting for prevalent glaucoma and myopia. GCC = ganglion cell complex, RNFL = retinal nerve fiber layer, GCL = ganglion cell layer, IPL = inner plexiform layer, INL = inner nuclear layer, OPL+ONL = outer plexiform layer plus outer nuclear layer, PS = photoreceptor segment layer, RPE+BM = retinal pigment epithelium plus Bruch's membrane, CSI = choroid scleral interface.

**a.**

**Association of retinal layer thickness with prevalent AMD**

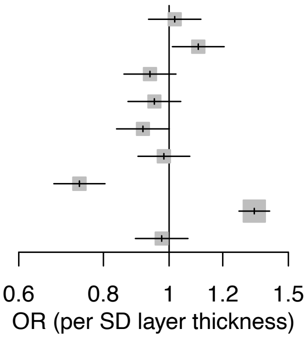


| | OR | 95% CI | P | Cases (N) | Controls (N) |
|---|---|---|---|---|---|
| GCC | 1.02 | [0.93; 1.11] | 0.67 | 512 | 44,311 |
| RNFL | 1.10 | [1.01; 1.21] | 0.027 | 512 | 44,311 |
| GCL | 0.94 | [0.86; 1.02] | 0.15 | 512 | 44,311 |
| IPL | 0.95 | [0.87; 1.04] | 0.27 | 512 | 44,311 |
| INL | 0.91 | [0.84; 1.00] | 0.052 | 512 | 44,311 |
| OPL+ONL | 0.98 | [0.90; 1.07] | 0.69 | 512 | 44,311 |
| PS | 0.74 | [0.68; 0.80] | 6.6e−12 | 512 | 44,311 |
| RPE+BM | 1.34 | [1.27; 1.41] | 1.3e−27 | 512 | 44,311 |
| CSI | 0.98 | [0.89; 1.07] | 0.58 | 512 | 44,311 |

**b.**

**Association of retinal layer thickness with incident AMD**

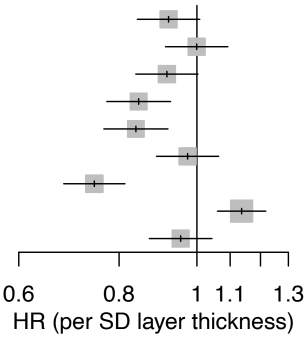


| | HR | 95% CI | P | Cases (N) | Controls (N) |
|---|---|---|---|---|---|
| GCC | 0.92 | [0.84; 1.01] | 0.075 | 494 | 43,817 |
| RNFL | 1.00 | [0.91; 1.09] | 0.99 | 494 | 43,817 |
| GCL | 0.92 | [0.84; 1.00] | 0.058 | 494 | 43,817 |
| IPL | 0.85 | [0.77; 0.93] | 0.00034 | 494 | 43,817 |
| INL | 0.84 | [0.77; 0.92] | 0.00019 | 494 | 43,817 |
| OPL+ONL | 0.97 | [0.89; 1.06] | 0.56 | 494 | 43,817 |
| PS | 0.75 | [0.68; 0.81] | 4.9e−11 | 494 | 43,817 |
| RPE+BM | 1.14 | [1.06; 1.22] | 0.00029 | 494 | 43,817 |
| CSI | 0.95 | [0.87; 1.04] | 0.31 | 494 | 43,817 |

Supplementary Figure 6: Association of retinal layer thicknesses with prevalent and incident AMD, additionally adjusting for prevalent glaucoma and myopia. This is the correlate to Figure 2, additionally adjusted for glaucoma and myopia at time of image acquisition. GCC = ganglion cell complex, RNFL = retinal nerve fiber layer, GCL = ganglion cell layer, IPL = inner plexiform layer, INL = inner nuclear layer, OPL+ONL = outer plexiform layer plus outer nuclear layer, PS = photoreceptor segment layer, RPE+BM = retinal pigment epithelium plus Bruch's membrane, CSI = choroid scleral interface.

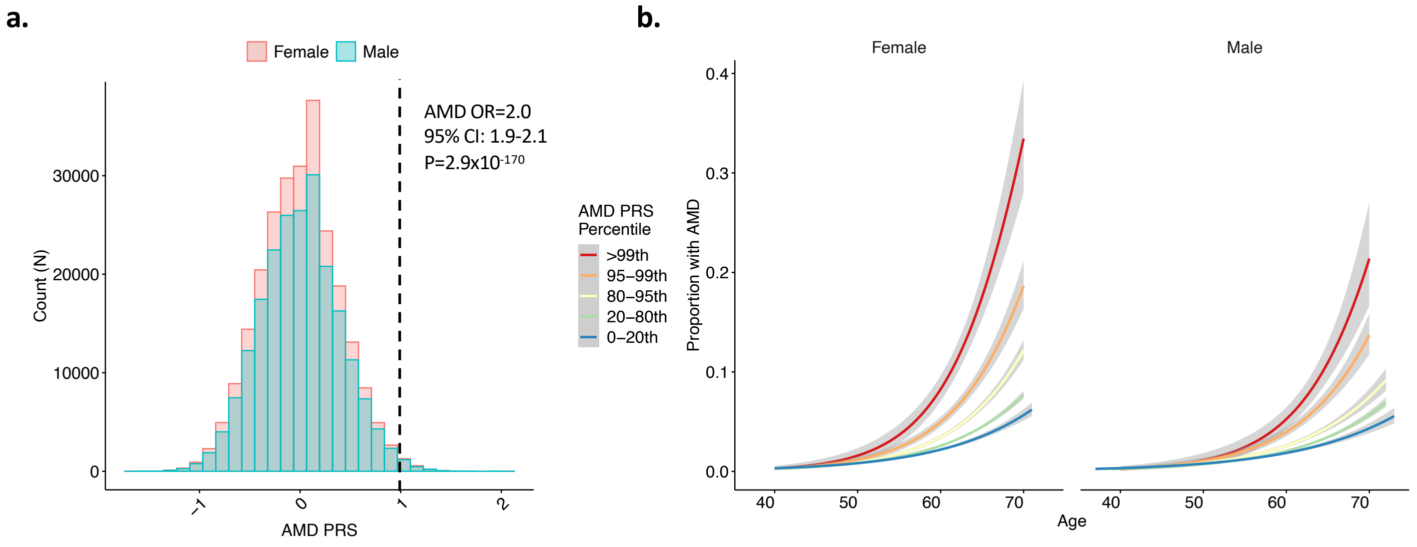


Supplementary Figure 7: Association of AMD PRS with AMD stratified by age. (a) Distribution of AMD PRS in females and males, whereby 1 unit increase in the AMD PRS confers a 2-fold increased risk of AMD (95%CI 1.9-2.1, $P=2.9\times10^{-170}$). (b) Association of age with proportion of individuals with prevalent or incident AMD, stratified by AMD PRS percentile and sex. Curves and standard errors reflect the best-fit generalized additive model (gam) to the individual-level data, with added smoothness. AMD = age-related macular degeneration; PRS = polygenic risk score.

**a.**

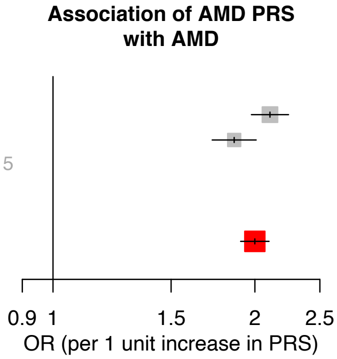


| | OR | 95% CI | P | Cases (N) | Controls (N) |
|---|---|---|---|---|---|
| Female | 2.11 | [1.98; 2.25] | 7.2e−116 | 6,306 | 235,371 |
| Male | 1.86 | [1.73; 2.01] | 1.3e−58 | 4,445 | 202,396 |
| Heterogeneity: *p* = 0.015 | | | | | |
| All | 2.00 | [1.90; 2.10] | 2.9e−170 | 10,751 | 437,767 |

**b.**

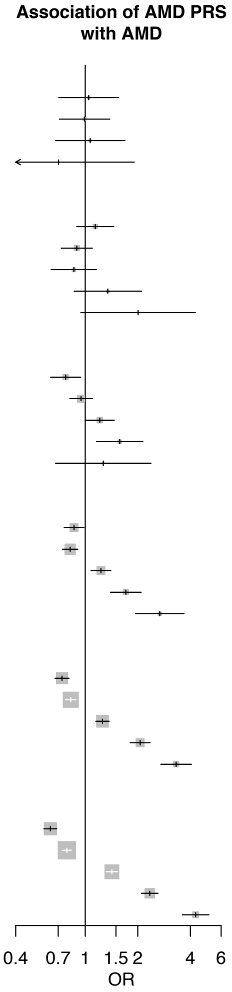


Age: / AMD PRS Percentile:

| | OR | 95% CI | P | Cases in PRS & Age Bin (N) | Controls in PRS & Age Bin (N) | All Cases in Age Bin (N) | All Controls in Age Bin (N) |
|---|---|---|---|---|---|---|---|
| **[40,45)** | | | | | | | |
| 0–20th | 1.05 | [0.71; 1.55] | 0.82 | 33 | 9,441 | 144 | 44,755 |
| 20–80th | 0.99 | [0.71; 1.38] | 0.96 | 84 | 26,145 | 144 | 44,755 |
| 80–95th | 1.07 | [0.67; 1.69] | 0.78 | 22 | 6,816 | 144 | 44,755 |
| 95–99th | 0.70 | [0.26; 1.91] | 0.49 | 4 | 1,880 | 144 | 44,755 |
| **[45,50)** | | | | | | | |
| 0–20th | 1.14 | [0.89; 1.46] | 0.29 | 86 | 12,226 | 374 | 58,598 |
| 20–80th | 0.89 | [0.73; 1.10] | 0.29 | 210 | 34,468 | 374 | 58,598 |
| 80–95th | 0.86 | [0.64; 1.16] | 0.33 | 50 | 8,888 | 374 | 58,598 |
| 95–99th | 1.35 | [0.86; 2.10] | 0.19 | 21 | 2,470 | 374 | 58,598 |
| >99th | 2.01 | [0.94; 4.27] | 0.07 | 7 | 546 | 374 | 58,598 |
| **[50,55)** | | | | | | | |
| 0–20th | 0.77 | [0.63; 0.94] | 0.011 | 120 | 13,840 | 717 | 68,073 |
| 20–80th | 0.95 | [0.82; 1.10] | 0.47 | 411 | 39,956 | 717 | 68,073 |
| 80–95th | 1.21 | [1.00; 1.47] | 0.05 | 131 | 10,716 | 717 | 68,073 |
| 95–99th | 1.58 | [1.16; 2.14] | 0.0035 | 45 | 2,807 | 717 | 68,073 |
| >99th | 1.27 | [0.68; 2.38] | 0.46 | 10 | 754 | 717 | 68,073 |
| **[55,60)** | | | | | | | |
| 0–20th | 0.86 | [0.76; 0.98] | 0.027 | 286 | 16,707 | 1,552 | 80,326 |
| 20–80th | 0.82 | [0.74; 0.91] | 0.00012 | 838 | 47,331 | 1,552 | 80,326 |
| 80–95th | 1.23 | [1.08; 1.40] | 0.0019 | 283 | 12,262 | 1,552 | 80,326 |
| 95–99th | 1.71 | [1.39; 2.09] | 2.4e−07 | 104 | 3,225 | 1,552 | 80,326 |
| >99th | 2.67 | [1.94; 3.68] | 1.6e−09 | 41 | 801 | 1,552 | 80,326 |
| **[60,65)** | | | | | | | |
| 0–20th | 0.74 | [0.67; 0.81] | 1.3e−10 | 547 | 21,862 | 3,389 | 105,953 |
| 20–80th | 0.83 | [0.77; 0.89] | 5.6e−08 | 1,842 | 62,513 | 3,389 | 105,953 |
| 80–95th | 1.26 | [1.15; 1.37] | 4.5e−07 | 631 | 16,399 | 3,389 | 105,953 |
| 95–99th | 2.06 | [1.81; 2.35] | 2e−27 | 261 | 4,131 | 3,389 | 105,953 |
| >99th | 3.32 | [2.71; 4.06] | 2.9e−31 | 108 | 1,048 | 3,389 | 105,953 |
| **[65,70]** | | | | | | | |
| 0–20th | 0.63 | [0.58; 0.69] | 2.5e−26 | 658 | 16,721 | 4,575 | 80,062 |
| 20–80th | 0.79 | [0.74; 0.83] | 2.4e−15 | 2,429 | 47,280 | 4,575 | 80,062 |
| 80–95th | 1.42 | [1.32; 1.53] | 3.6e−20 | 933 | 12,242 | 4,575 | 80,062 |
| 95–99th | 2.34 | [2.10; 2.61] | 4.8e−52 | 395 | 3,140 | 4,575 | 80,062 |
| >99th | 4.29 | [3.60; 5.11] | 4.8e−59 | 160 | 679 | 4,575 | 80,062 |

Supplementary Figure 8: Association of AMD PRS with AMD stratified by sex and age. (a) Association of AMD PRS with AMD by sex shows significant heterogeneity ($P_{heterogeneity}$=0.015), with females having higher odds of AMD per unit increase in AMD PRS compared to males. Analyses by sex are adjusted for age, age$^2$, smoking status, and principal components of genetic ancestry. AMD = age-related macular degeneration; PRS = polygenic risk score. (b) association of AMD PRS percentile with AMD among 5 year age categories. Age-stratified analyses are adjusted for sex, smoking status, and principal components of genetic ancestry. AMD = age-related macular degeneration; PRS = polygenic risk score.

**a.**

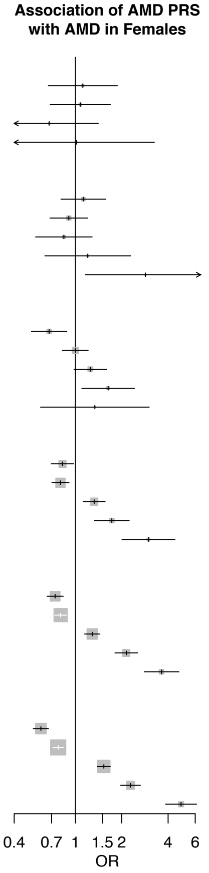


| Age bin / PRS percentile | OR | 95% CI | P | Cases in PRS & Age Bin (N) | Controls in PRS & Age Bin (N) | All Cases in Age Bin (N) | All Controls in Age Bin (N) |
|---|---|---|---|---|---|---|---|
| **[40,45)** | | | | | | | |
| 0–20th | 1.12 | [0.67; 1.87] | 0.67 | 20 | 5,153 | 79 | 23,743 |
| 20–80th | 1.08 | [0.69; 1.69] | 0.75 | 47 | 13,732 | 79 | 23,743 |
| 80–95th | 0.67 | [0.32; 1.41] | 0.29 | 8 | 3,629 | 79 | 23,743 |
| 95–99th | 1.02 | [0.32; 3.26] | 0.98 | 3 | 984 | 79 | 23,743 |
| **[45,50)** | | | | | | | |
| 0–20th | 1.13 | [0.81; 1.57] | 0.49 | 46 | 6,842 | 199 | 32,361 |
| 20–80th | 0.91 | [0.68; 1.20] | 0.49 | 113 | 19,063 | 199 | 32,361 |
| 80–95th | 0.84 | [0.55; 1.28] | 0.42 | 25 | 4,779 | 199 | 32,361 |
| 95–99th | 1.20 | [0.63; 2.29] | 0.57 | 10 | 1,383 | 199 | 32,361 |
| >99th | 2.85 | [1.16; 7.00] | 0.023 | 5 | 294 | 199 | 32,361 |
| **[50,55)** | | | | | | | |
| 0–20th | 0.67 | [0.52; 0.88] | 0.0035 | 67 | 7,808 | 437 | 38,322 |
| 20–80th | 1.00 | [0.82; 1.21] | 0.98 | 256 | 22,488 | 437 | 38,322 |
| 80–95th | 1.25 | [0.98; 1.60] | 0.074 | 81 | 6,071 | 437 | 38,322 |
| 95–99th | 1.63 | [1.10; 2.42] | 0.015 | 27 | 1,542 | 437 | 38,322 |
| >99th | 1.34 | [0.59; 3.02] | 0.48 | 6 | 413 | 437 | 38,322 |
| **[55,60)** | | | | | | | |
| 0–20th | 0.82 | [0.70; 0.97] | 0.024 | 172 | 9,296 | 957 | 44,431 |
| 20–80th | 0.80 | [0.70; 0.91] | 0.00063 | 510 | 26,157 | 957 | 44,431 |
| 80–95th | 1.32 | [1.12; 1.56] | 0.0008 | 184 | 6,778 | 957 | 44,431 |
| 95–99th | 1.72 | [1.33; 2.23] | 3.9e−05 | 64 | 1,775 | 957 | 44,431 |
| >99th | 2.98 | [2.01; 4.43] | 6.4e−08 | 27 | 425 | 957 | 44,431 |
| **[60,65)** | | | | | | | |
| 0–20th | 0.74 | [0.65; 0.83] | 9.9e−07 | 322 | 11,718 | 1,991 | 56,984 |
| 20–80th | 0.80 | [0.73; 0.88] | 1.4e−06 | 1,073 | 33,803 | 1,991 | 56,984 |
| 80–95th | 1.29 | [1.15; 1.44] | 2e−05 | 373 | 8,717 | 1,991 | 56,984 |
| 95–99th | 2.14 | [1.81; 2.54] | 1.6e−18 | 156 | 2,196 | 1,991 | 56,984 |
| >99th | 3.63 | [2.80; 4.70] | 1.7e−22 | 67 | 550 | 1,991 | 56,984 |
| **[65,70)** | | | | | | | |
| 0–20th | 0.60 | [0.53; 0.67] | 4.9e−19 | 367 | 8,391 | 2,643 | 39,530 |
| 20–80th | 0.77 | [0.71; 0.84] | 1.6e−10 | 1,392 | 23,318 | 2,643 | 39,530 |
| 80–95th | 1.53 | [1.39; 1.69] | 1.2e−17 | 568 | 6,006 | 2,643 | 39,530 |
| 95–99th | 2.28 | [1.96; 2.64] | 1.6e−27 | 218 | 1,505 | 2,643 | 39,530 |
| >99th | 4.86 | [3.85; 6.12] | 1.2e−40 | 98 | 310 | 2,643 | 39,530 |

**b.**

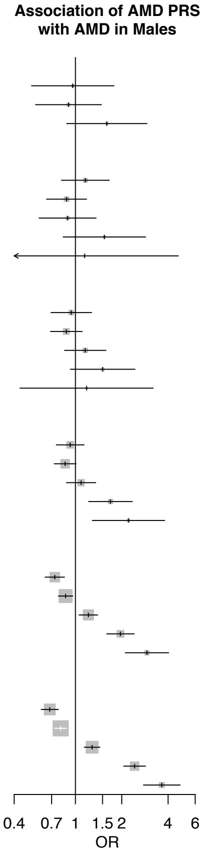


| Age bin / PRS percentile | OR | 95% CI | P | Cases in PRS & Age Bin (N) | Controls in PRS & Age Bin (N) | All Cases in Age Bin (N) | All Controls in Age Bin (N) |
|---|---|---|---|---|---|---|---|
| **[40,45)** | | | | | | | |
| 0–20th | 0.96 | [0.52; 1.78] | 0.9 | 13 | 4,288 | 65 | 21,012 |
| 20–80th | 0.90 | [0.55; 1.48] | 0.68 | 37 | 12,413 | 65 | 21,012 |
| 80–95th | 1.60 | [0.88; 2.92] | 0.12 | 14 | 3,187 | 65 | 21,012 |
| **[45,50)** | | | | | | | |
| 0–20th | 1.16 | [0.81; 1.66] | 0.41 | 40 | 5,384 | 175 | 26,237 |
| 20–80th | 0.88 | [0.65; 1.18] | 0.39 | 97 | 15,405 | 175 | 26,237 |
| 80–95th | 0.89 | [0.58; 1.36] | 0.59 | 25 | 4,109 | 175 | 26,237 |
| 95–99th | 1.54 | [0.83; 2.86] | 0.17 | 11 | 1,087 | 175 | 26,237 |
| >99th | 1.15 | [0.28; 4.66] | 0.85 | 2 | 252 | 175 | 26,237 |
| **[50,55)** | | | | | | | |
| 0–20th | 0.94 | [0.69; 1.27] | 0.69 | 53 | 6,032 | 280 | 29,751 |
| 20–80th | 0.87 | [0.69; 1.11] | 0.26 | 155 | 17,468 | 280 | 29,751 |
| 80–95th | 1.16 | [0.85; 1.58] | 0.35 | 50 | 4,645 | 280 | 29,751 |
| 95–99th | 1.50 | [0.93; 2.44] | 0.098 | 18 | 1,265 | 280 | 29,751 |
| >99th | 1.18 | [0.44; 3.20] | 0.74 | 4 | 341 | 280 | 29,751 |
| **[55,60)** | | | | | | | |
| 0–20th | 0.92 | [0.75; 1.14] | 0.46 | 114 | 7,411 | 595 | 35,895 |
| 20–80th | 0.86 | [0.73; 1.01] | 0.063 | 328 | 21,174 | 595 | 35,895 |
| 80–95th | 1.09 | [0.87; 1.35] | 0.45 | 99 | 5,484 | 595 | 35,895 |
| 95–99th | 1.69 | [1.22; 2.34] | 0.0017 | 40 | 1,450 | 595 | 35,895 |
| >99th | 2.21 | [1.29; 3.80] | 0.0041 | 14 | 376 | 595 | 35,895 |
| **[60,65)** | | | | | | | |
| 0–20th | 0.73 | [0.64; 0.85] | 3.2e−05 | 225 | 10,144 | 1,398 | 48,969 |
| 20–80th | 0.86 | [0.78; 0.96] | 0.0071 | 769 | 28,710 | 1,398 | 48,969 |
| 80–95th | 1.21 | [1.06; 1.39] | 0.0059 | 258 | 7,682 | 1,398 | 48,969 |
| 95–99th | 1.96 | [1.60; 2.41] | 1.3e−10 | 105 | 1,935 | 1,398 | 48,969 |
| >99th | 2.92 | [2.11; 4.03] | 1e−10 | 41 | 498 | 1,398 | 48,969 |
| **[65,70)** | | | | | | | |
| 0–20th | 0.68 | [0.60; 0.77] | 3.8e−09 | 291 | 8,330 | 1,932 | 40,532 |
| 20–80th | 0.80 | [0.73; 0.88] | 2.3e−06 | 1,037 | 23,962 | 1,932 | 40,532 |
| 80–95th | 1.28 | [1.14; 1.44] | 3.3e−05 | 365 | 6,236 | 1,932 | 40,532 |
| 95–99th | 2.42 | [2.06; 2.85] | 2.4e−26 | 177 | 1,635 | 1,932 | 40,532 |
| >99th | 3.64 | [2.77; 4.79] | 2.5e−20 | 62 | 369 | 1,932 | 40,532 |

Supplementary Figure 9: Association of AMD PRS with AMD in (a) Females and (b) Males, by 5-year age groups. Associations with AMD are provided for each PRS percentile within that age group, in a logistic regression model adjusted for smoking status and principal components of genetic ancestry. AMD = age-related macular degeneration, PRS = polygenic risk score.

**a.**

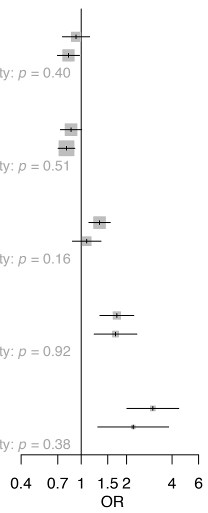


| Association of AMD PRS with AMD among Age [55,60) | OR | 95% CI | P | Cases in PRS & Age Bin (N) | Controls in PRS & Age Bin (N) | All Cases in Age Bin (N) | All Controls in Age Bin (N) |
|---|---|---|---|---|---|---|---|
| **0–20th** | | | | | | | |
| Male | 0.92 | [0.75; 1.14] | 0.46 | 114 | 7,411 | 595 | 35,895 |
| Female | 0.82 | [0.70; 0.97] | 0.024 | 172 | 9,296 | 957 | 44,431 |
| Heterogeneity: *p* = 0.40 | | | | | | | |
| **20–80th** | | | | | | | |
| Male | 0.86 | [0.73; 1.01] | 0.063 | 328 | 21,174 | 595 | 35,895 |
| Female | 0.80 | [0.70; 0.91] | 0.00063 | 510 | 26,157 | 957 | 44,431 |
| Heterogeneity: *p* = 0.51 | | | | | | | |
| **80–95th** | | | | | | | |
| Female | 1.32 | [1.12; 1.56] | 0.0008 | 184 | 6,778 | 957 | 44,431 |
| Male | 1.09 | [0.87; 1.35] | 0.45 | 99 | 5,484 | 595 | 35,895 |
| Heterogeneity: *p* = 0.16 | | | | | | | |
| **95–99th** | | | | | | | |
| Female | 1.72 | [1.33; 2.23] | 3.9e–05 | 64 | 1,775 | 957 | 44,431 |
| Male | 1.69 | [1.22; 2.34] | 0.0017 | 40 | 1,450 | 595 | 35,895 |
| Heterogeneity: *p* = 0.92 | | | | | | | |
| **>99th** | | | | | | | |
| Female | 2.98 | [2.01; 4.43] | 6.4e–08 | 27 | 425 | 957 | 44,431 |
| Male | 2.21 | [1.29; 3.80] | 0.0041 | 14 | 376 | 595 | 35,895 |
| Heterogeneity: *p* = 0.38 | | | | | | | |

**b.**

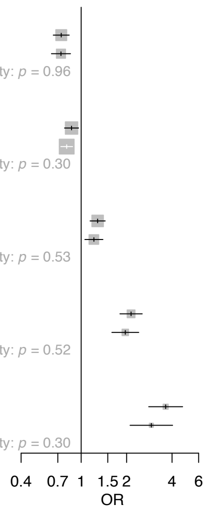


| Association of AMD PRS with AMD among Age [60,65) | OR | 95% CI | P | Cases in PRS & Age Bin (N) | Controls in PRS & Age Bin (N) | All Cases in Age Bin (N) | All Controls in Age Bin (N) |
|---|---|---|---|---|---|---|---|
| **0–20th** | | | | | | | |
| Female | 0.74 | [0.65; 0.83] | 9.9e–07 | 322 | 11,718 | 1,991 | 56,984 |
| Male | 0.73 | [0.64; 0.85] | 3.2e–05 | 225 | 10,144 | 1,398 | 48,969 |
| Heterogeneity: *p* = 0.96 | | | | | | | |
| **20–80th** | | | | | | | |
| Male | 0.86 | [0.78; 0.96] | 0.0071 | 769 | 28,710 | 1,398 | 48,969 |
| Female | 0.80 | [0.73; 0.88] | 1.4e–06 | 1,073 | 33,803 | 1,991 | 56,984 |
| Heterogeneity: *p* = 0.30 | | | | | | | |
| **80–95th** | | | | | | | |
| Female | 1.29 | [1.15; 1.44] | 2e–05 | 373 | 8,717 | 1,991 | 56,984 |
| Male | 1.21 | [1.06; 1.39] | 0.0059 | 258 | 7,682 | 1,398 | 48,969 |
| Heterogeneity: *p* = 0.53 | | | | | | | |
| **95–99th** | | | | | | | |
| Female | 2.14 | [1.81; 2.54] | 1.6e–18 | 156 | 2,196 | 1,991 | 56,984 |
| Male | 1.96 | [1.60; 2.41] | 1.3e–10 | 105 | 1,935 | 1,398 | 48,969 |
| Heterogeneity: *p* = 0.52 | | | | | | | |
| **>99th** | | | | | | | |
| Female | 3.63 | [2.80; 4.70] | 1.7e–22 | 67 | 550 | 1,991 | 56,984 |
| Male | 2.92 | [2.11; 4.03] | 1e–10 | 41 | 498 | 1,398 | 48,969 |
| Heterogeneity: *p* = 0.30 | | | | | | | |

**c.**

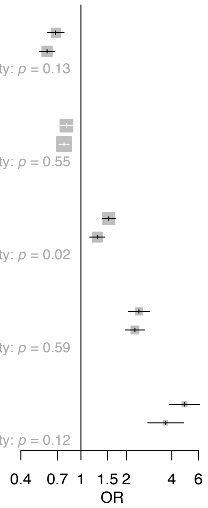


| Association of AMD PRS with AMD among Age [65,70) | OR | 95% CI | P | Cases in PRS & Age Bin (N) | Controls in PRS & Age Bin (N) | All Cases in Age Bin (N) | All Controls in Age Bin (N) |
|---|---|---|---|---|---|---|---|
| **0–20th** | | | | | | | |
| Male | 0.68 | [0.60; 0.77] | 3.8e–09 | 291 | 8,330 | 1,932 | 40,532 |
| Female | 0.60 | [0.53; 0.67] | 4.9e–19 | 367 | 8,391 | 2,643 | 39,530 |
| Heterogeneity: *p* = 0.13 | | | | | | | |
| **20–80th** | | | | | | | |
| Male | 0.80 | [0.73; 0.88] | 2.3e–06 | 1,037 | 23,962 | 1,932 | 40,532 |
| Female | 0.77 | [0.71; 0.84] | 1.6e–10 | 1,392 | 23,318 | 2,643 | 39,530 |
| Heterogeneity: *p* = 0.55 | | | | | | | |
| **80–95th** | | | | | | | |
| Female | 1.53 | [1.39; 1.69] | 1.2e–17 | 568 | 6,006 | 2,643 | 39,530 |
| Male | 1.28 | [1.14; 1.44] | 3.3e–05 | 365 | 6,236 | 1,932 | 40,532 |
| Heterogeneity: *p* = 0.02 | | | | | | | |
| **95–99th** | | | | | | | |
| Male | 2.42 | [2.06; 2.85] | 2.4e–26 | 177 | 1,635 | 1,932 | 40,532 |
| Female | 2.28 | [1.96; 2.64] | 1.6e–27 | 218 | 1,505 | 2,643 | 39,530 |
| Heterogeneity: *p* = 0.59 | | | | | | | |
| **>99th** | | | | | | | |
| Female | 4.86 | [3.85; 6.12] | 1.2e–40 | 98 | 310 | 2,643 | 39,530 |
| Male | 3.64 | [2.77; 4.79] | 2.5e–20 | 62 | 369 | 1,932 | 40,532 |
| Heterogeneity: *p* = 0.12 | | | | | | | |

Supplementary Figure 10: Heterogeneity of association of the AMD PRS with AMD by sex among different age groups. Associations with AMD are provided for each PRS percentile within individuals between ages (a) 55-60, (b) 60-65, and (c) 65-70, in a logistic regression model adjusted for smoking status and principal components of genetic ancestry. AMD = age-related macular degeneration, PRS = polygenic risk score.

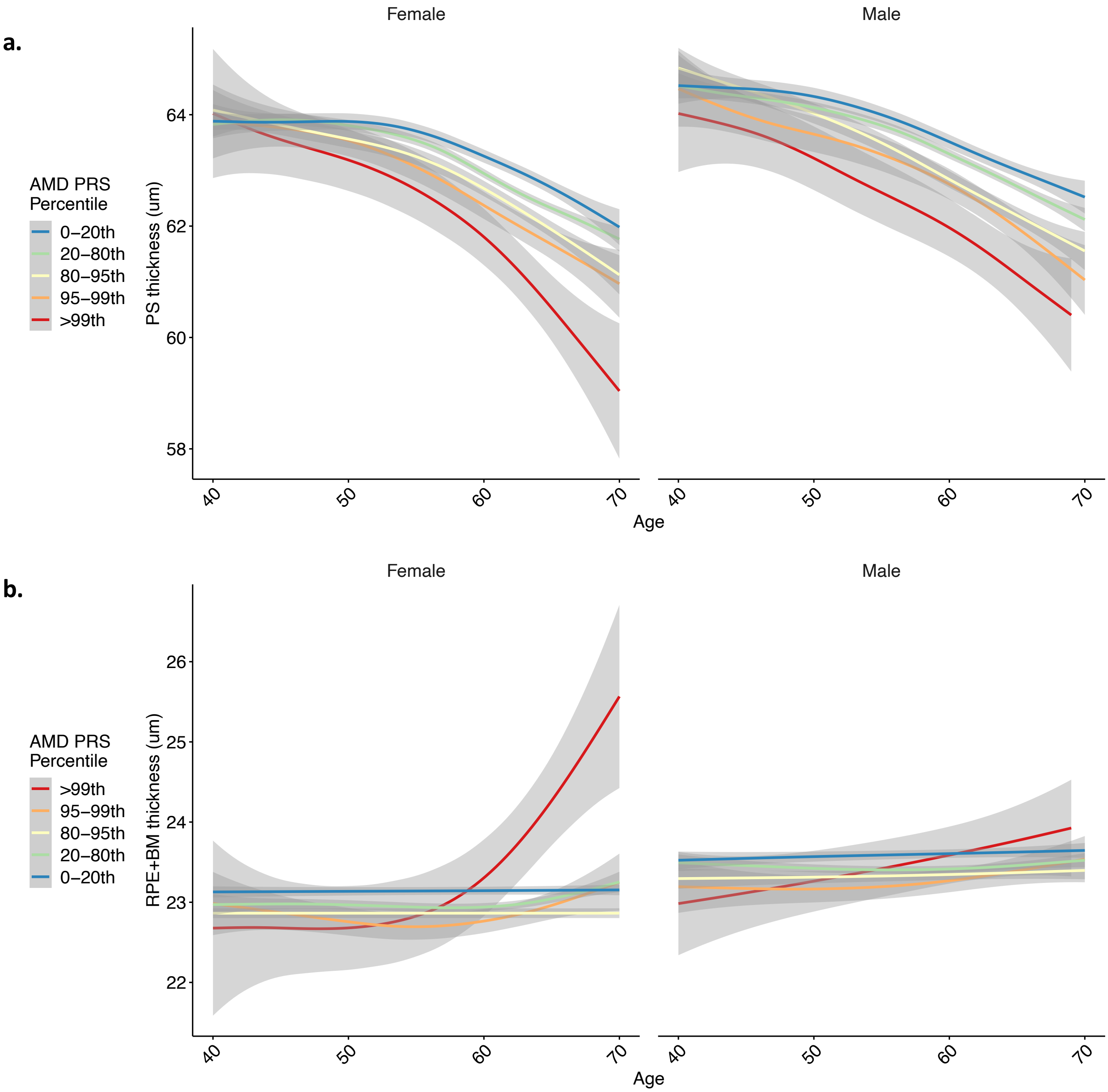


Supplementary Figure 11: Relationship of age with (a) photoreceptor and (b) RPE+BM layer thicknesses by sex and AMD PRS percentile. Curves and standard errors in panels (a) and (b) reflect the best-fit generalized additive model (gam) to the individual-level data, with added smoothness. PRS = polygenic risk score; PS = photoreceptor segment layer; RPE+BM = retinal pigment epithelium plus Bruch’s membrane complex.

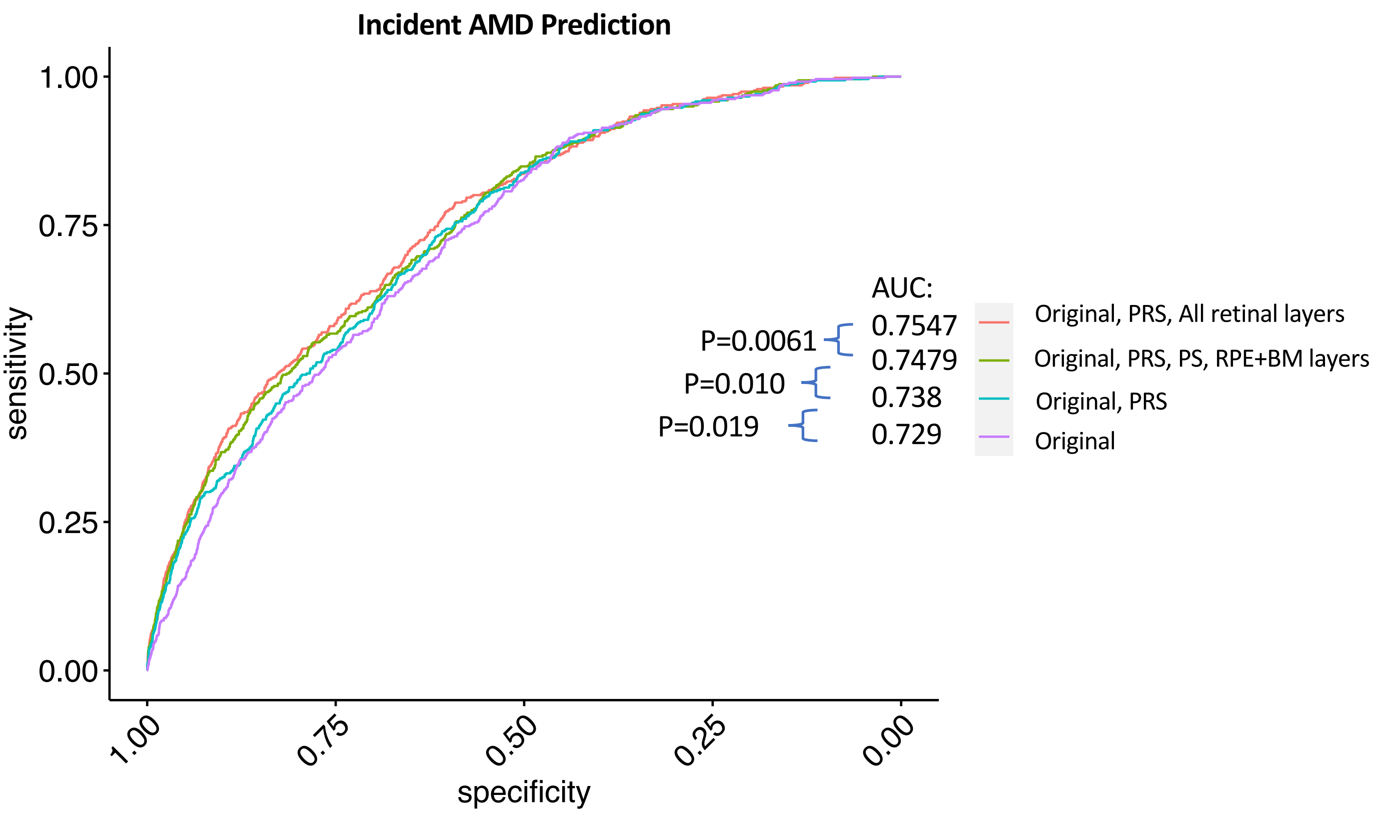


Supplementary Figure 12: Prediction of incident AMD significantly improves with addition of retinal layer thicknesses from OCT, on top of baseline characteristics and a polygenetic risk score. Prediction of incident AMD using a cox model where Original refers to the following exposures: age, $age^2$, sex, smoking status, BMI, and the first ten principal components of genetic ancestry**.** A tiered approach successively adding on a polygenic risk score (PRS), PS and RPE+BM layer thicknesses, and lastly thicknesses of all retinal layers, shows successive significant improvement in AUC. AUC=area under the curve. PRS=polygenic risk score, PS=photoreceptor segment thickness, RPE+BM=retinal pigment epithelium and Bruch's membrane thickness.
